# Pharmacogenomic study of heart failure and candesartan response from the CHARM programme

**DOI:** 10.1101/2021.09.28.21263908

**Authors:** Marie-Pierre Dubé, Olympe Chazara, Audrey Lemaçon, Géraldine Asselin, Sylvie Provost, Amina Barhdadi, Louis-Philippe Lemieux Perreault, Ian Mongrain, Quanli Wang, Keren Carss, Dirk S Paul, Jonathan W Cunningham, Jean Rouleau, Scott D Solomon, John J V McMurray, Salim Yusuf, Chris B Granger, Carolina Haefliger, Simon de Denus, Jean-Claude Tardif

## Abstract

**Aims:** The Candesartan in Heart failure Assessment of Reduction in Mortality and morbidity (CHARM) programme consisted of three parallel, randomised, double-blind clinical trials comparing candesartan with placebo in patients with heart failure (HF) categorised according to left ventricular ejection fraction and tolerability to an ACE inhibitor. We conducted a pharmacogenomic study of the CHARM studies to identify genetic predictors of heart failure progression and the efficacy and safety of treatment with candesartan.

**Methods:** We performed genome-wide association studies (GWAS) with the composite endpoint of cardiovascular death or hospitalisation for heart failure in 2727 patients from CHARM-Overall and stratified by CHARM study according to preserved and reduced ejection fraction. The safety endpoints were hyperkalaemia, renal dysfunction, hypotension, and change in systolic blood pressure. We also conducted a genome-wide gene-level collapsing analysis from whole-exome sequencing data with the composite cardiovascular endpoint.

**Results:** We found the genetic variant rs66886237 at 8p21.3 near the gene *GFRA2* to be associated with the composite cardiovascular endpoint in 1029 HF patients with preserved ejection fraction from the CHARM-Preserved study [hazard ratio (HR): 1.91, 95% confidence interval (CI): 1.55-2.35; P=1.7×10^-9^], but not in patients with reduced ejection fraction. None of the GWAS for candesartan safety or efficacy passed the significance threshold.

**Conclusions:** We have identified a candidate genetic variant potentially predictive of the progression of heart failure in patients with preserved ejection fraction. The findings require further replication and we cannot exclude the possibility that the results may be chance findings.

## INTRODUCTION

Candesartan is an angiotensin II receptor blocker (ARB) that is widely used alone or in combination with other agents as therapy for hypertension and heart failure (HF). Multiple mechanistic studies alluded to the potential benefits of inhibition of the renin-angiotensin-aldosterone system (RAAS) with ARBs in HF,^1-3^ which were subsequently confirmed in large clinical trials, although this benefit varied depending on the population studied and the concomitant medication used.^4-7^

Candesartan is a selective AT1 subtype angiotensin II receptor antagonist, which is orally administered as candesartan cilexetil, a prodrug, which undergoes hydrolysis to candesartan during absorption from the gastrointestinal tract. Candesartan is not significantly metabolised by the cytochrome P450 system and at therapeutic concentrations has no effects on P450 enzymes. Candesartan was shown to be beneficial in patients with heart failure in the Candesartan in Heart failure Assessment of Reduction in Mortality and morbidity (CHARM) programme designed as three parallel, independent, integrated, randomised, double-blind clinical trials comparing candesartan with placebo in three distinct populations of patients with NYHA class II–IV HF based on participants’ assessment of left ventricular ejection fraction (LVEF) and history of tolerability to an ACE inhibitor.^8^ The primary endpoint of each trial was testing whether the use of candesartan would reduce the risk of cardiovascular death or hospital admission for HF. In CHARM-Alternative and CHARM-Added, the primary endpoint was significantly reduced with candesartan as compared to placebo (HR 0.77, 95% CI 0.67-0.89, p= 0.0004; and HR 0.85, 95% CI 0.75-0.96, p= 0.011, respectively). In CHARM-Preserved, however, the primary composite endpoint did not reach significance (HR= 0.89, 95% CI 0.77–1.03. P= 0.118). CHARM-Overall showed a highly significant reduction in the combined incidence of cardiovascular death and hospitalisation for HF (HR= 0.84, 95% CI 0.77–0.91, p<0.0001).

Here, we present a post-hoc pharmacogenomic study of the CHARM programme in a subgroup of the original study participants, with the aim of identifying genetic predictors of the progression of HF and for the efficacy and safety of treatment with candesartan.

## METHODS

### Study data

The CHARM programme was designed as three parallel, independent, integrated, randomised, double-blind clinical trials comparing candesartan with placebo in three distinct populations of patients with NYHA class II–IV HF based on participants’ assessment of left ventricular ejection fraction (LVEF) and history of tolerability to an ACE inhibitor.^8^ CHARM-Alternative included patients with depressed LV function (EF ≤ 40%) and who were not treated with ACE inhibitors due to intolerance,^4^ CHARM-Added included 2548 patients with depressed LV function (EF ≤ 40%) and treated with ACE inhibitors,^5^ and CHARM-Preserved included 3025 patients with preserved LV function (EF > 40%) with or without ACE inhibitors.^6^ CHARM-Overall was the joint analysis of all 3 studies combined.^9^ The CHARM studies involved 26 countries and 618 sites. The first patient was randomised on March 22 1999 and the last patient completed the study on March 31 2003, with a median follow-up time of 37.8 months. The active treatment group received candesartan once daily, at a starting dose of 4 mg or 8 mg once daily, which was up-titrated by doubling the dose at 2-week intervals to a maximum of 32 mg once daily or the highest tolerated level. Main exclusion criteria were serum creatinine of 265 μmol/L or more, serum potassium of 5.5 mmol/L or more, bilateral renal artery stenosis, symptomatic hypotension, critical aortic or mitral stenosis, myocardial infarction, stroke, or open-heart surgery in the previous 4 weeks, use of an angiotensin-receptor blocker in the previous 2 weeks. Other exclusion criteria have been previously described.^8^

In this pharmacogenetic sub-study, there were 2727 participants in CHARM-Overall, which included 755 participants from CHARM-Alternative, 943 from CHARM-Added, and 1029 from CHARM-Preserved (**Figure 1**).

**Figure 1.**
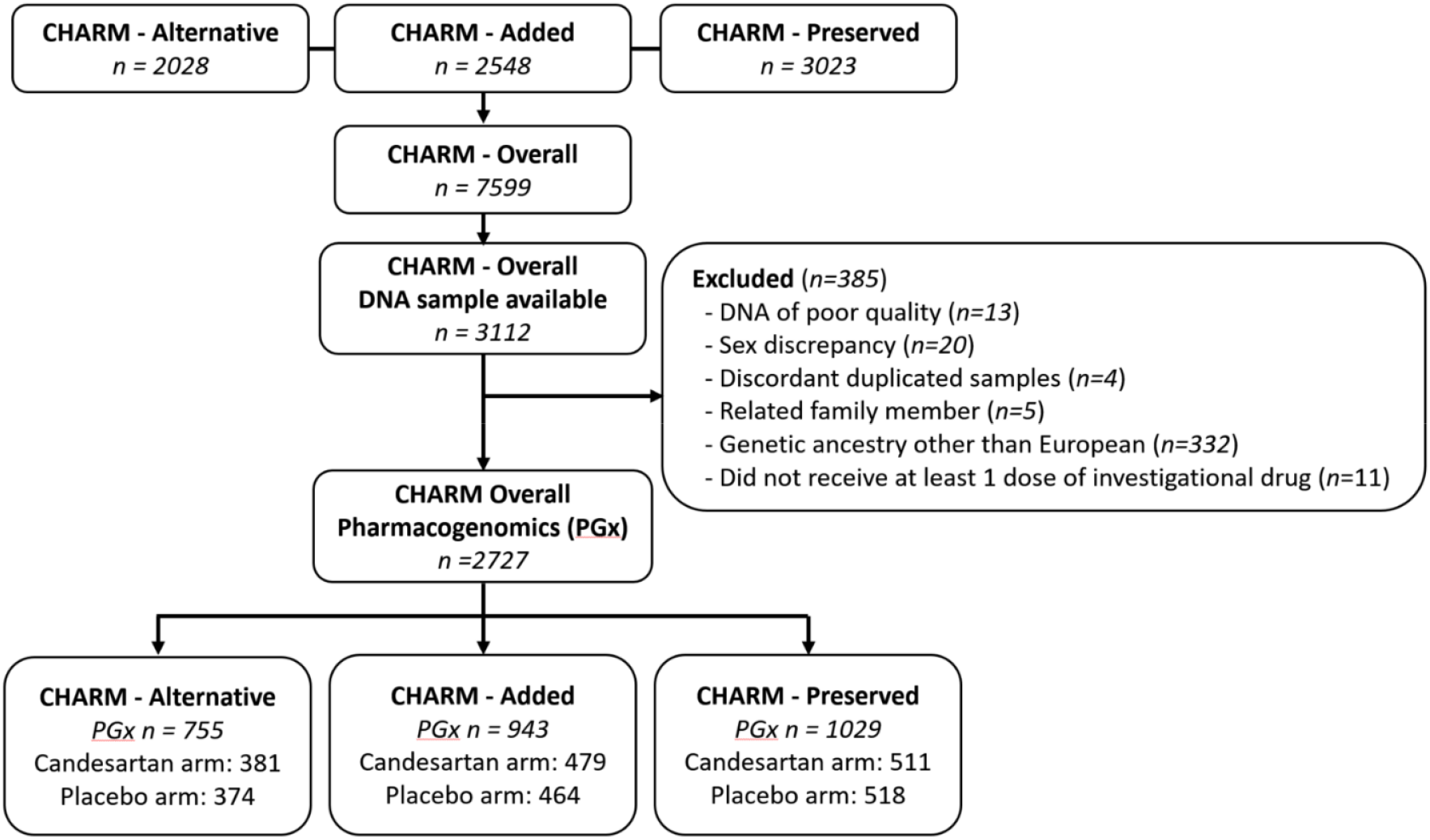
Flow diagram of participants of the CHARM pharmacogenomic study.

### Endpoint Definitions

The CV efficacy endpoint for the present pharmacogenomic study was defined identically to the primary endpoint of the individual CHARM studies and consists of a composite of CV death or hospitalisation for the management of chronic HF, whichever occurred first. Potential study endpoints were adjudicated by an independent clinical endpoint committee in the CHARM studies. Event-free patients who completed the study were censored at the date of study completion, and those who did not complete the study were censored at the date of last contact. Patients who died from a non-CV cause were censored at the time of death. The pharmacogenomic safety endpoints considered were hyperkalaemia, renal dysfunction, hypotension, and change in systolic blood pressure. Adverse events were recorded and encoded centrally. Systolic blood pressure was obtained during the physical examination at the scheduled 6-week visit and reported by investigators. Patients were followed up until study completion or date of last contact whichever occurred first.

### Genotyping

Genome-wide genotyping was performed using 200 ng of genomic DNA extracted from whole blood at the Beaulieu-Saucier Pharmacogenomics Centre (Montreal, Canada). The Illumina Infinium Multi-Ethnic Global Array (MEGA) Consortium v2 BeadChip (Illumina, San Diego, CA) including 2,036,060 genomic markers was used and processed according to the manufacturer’s specifications. BeadChips were scanned using the Illumina iScan Reader and analysed using Illumina’s Beeline v1.0.37.0 with the data manifest MEGA_Consortium_v2_15070954_A2.bpm, with minor manual cluster adjustment for ADME genes and using a custom cluster file. The Beeline final report files were used to generate gender plots, LRR and BAF graphics. PyGenClean ^10^ version 1.8.2 and PLINK ^11^ version 1.07 were used for the quality checks (QC) and genetic data cleanup process. The genotyping experiment consisted of 34 plates of DNA samples. There was one control per hybridization run (corresponding to 2 plates), selected from NA19119, NA18980, NA19147 and NA12878 obtained from the NIGMS Human Genetic Cell Repository at the Coriell Institute for Medical Research. The pairwise concordance of Coriell samples ranged from 0.999989 to 0.9999996. The comparison of Coriell genotypes to expectation from the 1000 Genomes data provided concordance ranging from 0.9959 to 0.9977.

Stepwise results of the genetic quality controls procedures are presented in **Supplementary Table 1**. Duplicated SNPs were evaluated for concordance, completion rate, allele call and minor allele frequency (MAF). SNPs with different allele calls or different MAF were retained. Identical and concordant SNPs were merged. The completion rate threshold for genotypes and samples was set to 98%. SNPs with genotyping plate bias (based on the 96 well plates used to dilute DNA samples) were flagged but not removed as the effect of genetic ancestry could not be excluded. Pairwise identity-by-state (IBS) was used to conduct close familial relationship checks. We removed all but one member of related samples (IBS2*ratio > 0.80) based on a selection of uncorrelated SNPs (r^2^ < 0.1). The pairwise IBS matrix was used as a distance metric to identify cryptic relatedness among samples and sample outliers by multidimensional scaling (MDS). The first two MDS components of each subject were plotted including the genotypes of HapMap CEU, JPT-CHB, and YRI population data (keeping only founder individuals). Outliers from the main cluster overlapping the CEU reference samples (Utah residents with Northern and Western European ancestry from the CEPH collection) were removed according to k-nearest neighbour with a threshold of 1.9σ in PyGenClean (v1.8.2) (**Supplementary Figure 1**). Principal components were generated on the study samples only, and the scree plot and the cumulative explained variance were used to select the principal components to control for confounding due to population structure. ^12^

### Imputation

Genome-wide imputation was performed using IMPUTE2 (v2.3.2)^13^ and phasing was performed using the SHAPEIT2 algorithm (v.2r790).^14^ Strand alignment was solved by flipping non A/T and C/G SNPs and 158,140 ambiguous A/T and C/G SNPs were considered missing and were imputed. Imputation was performed based on 1,194,173 genetic variants using the phased 1000 Genomes reference data with singleton sites removed released on June 16, 2014 and which include samples from all populations (distributed through the IMPUTE2 web site). The pseudo-autosomal regions on the X chromosome were imputed separately from the rest of the chromosome. Internal cross-validation was performed with IMPUTE2 and provided a mean genotype concordance of 98.1%. Any missing genotypes at the genotyped SNPs were also imputed. A total of 11,871,586 genetic variants with imputation probability of 0.90 or greater and completion rate of 98% or greater were retained for analyses. For the genome-wide analysis, there were a total of 5,140,623 genetic variants with MAF ≥ 5%.

### Statistical analysis

Genome-wide association studies (GWAS) of common genetic variants (MAF ≥ 0.05) were conducted using Genipe version 1.3.1. Cox proportional hazards regression was used for the composite endpoint of CV death or hospitalisation for HF, logistic regression was used for the dichotomous safety endpoints, and linear regression was used for the endpoint of change in systolic blood pressure. Imputation dosage for genotypes was used with the 1-degree of freedom additive genetic test adjusted for age, sex, and the first 10 principal components to control for genetic ancestry, with the addition of candesartan treatment when both study arms were included in the analyses. For the endpoint of change in systolic blood pressure, sensitivity analyses including adjustment for baseline systolic blood pressure value were conducted and are presented in **Supplementary Online Material**. GWAS were conducted in the CHARM-Overall programme and also stratified according to whether studies included HF patients with reduced or preserved ejection fraction (CHARM-Alternative + CHARM-Added versus CHARM-Preserved). GWAS aimed at the identification of genetic predictors of the progression of HF were conducted using patients in both arms of the CHARM studies. The GWAS aimed at the identification of genetic determinants of the efficacy and safety of treatment with candesartan were conducted using patients randomly assigned to the candesartan arm only and the identified genetic variants were then assessed for their effect in the placebo group and for interaction effects with candesartan treatment. Exploratory GWAS testing for the interaction term between each genetic variant and the treatment arm are presented in the **Supplementary Online Material**. Sensitivity analyses with the identified genetic variant in the CHARM-Preserved study were conducted with additional adjustment for BMI, left ventricular ejection fraction, history of atrial fibrillation, and prior MI, diabetes to assess possible mediation pathways. All analyses are considered exploratory and hypothesis-generating. Overall, we conducted 18 primary analysis GWAS, which were supported by exploratory and sensitivity analysis GWAS (**Supplementary Table 2**). Each GWAS is assessed at a significance level of 2.8×10^-9^ to adjust for the multiple testing of genetic variants and for the 18 primary analyses. Results are reported with point estimates and 95% confidence intervals (CI) which are not adjusted for multiple comparisons. Statistical analyses (except for GWAS and functional annotation analyses) were conducted using SAS versions 9.3 and 9.4, and the top findings from the GWAS were reproduced using SAS. The proportionality of hazards assumption was confirmed for the leading variants identified by GWAS using the Supremum test based on martingale residuals.

### Whole-exome sequencing data analysis

Genomic DNA was also exome-sequenced at the Columbia Institute for Genomic Medicine. Sequence capture was performed using the IDT xGen Exome Research Panel v1.0 kit and sequenced on Illumina’s NovaSeq 6000 platform with 150 bp paired-end reads. Sequences were processed through the AstraZeneca’s Centre for Genomics Research bioinformatics pipeline using a custom-built Amazon Web Services (AWS) cloud compute platform running Illumina DRAGEN Bio-IT Platform Germline Pipeline version 3.0.7. The reads were aligned to the GRCh38 genome reference, followed by single-nucleotide variant (SNV) and indel calling. SNVs and indels were annotated using SnpEFF v4.34 against Ensembl Build 38.92. Mean sequence coverage was 90.45x, with on average 97.28% of the target bases in a given sample achieving at least 10x coverage of the Consensus Coding Sequence (CCDS release 22).

We analysed rare variants in 18,500 coding genes in samples obtained from CHARM individuals that passed strict sample-level quality control as follow: contamination levels <4% based on VerifyBamID, concordance between genetic and self-declared sex, ≥95% of CCDS r22 bases covered with ≥10-fold coverage, unrelated up to the third degree according to PI_HAT (PLINK v1.07), and of genetically-determined European ancestry (peddy_ancestry_prob ≥0.98). Rare variants were analysed with collapsing analyses as previously described,^15^ for variants with a minimum read depth of 10, located within the CCDS r22 or the 2 bp canonical splice sites within the introns, and for variants that passed quality score thresholds as described in ^16^. Eleven models defining sets of qualifying variants (QV) were used on the basis of the variants’ predicted functional consequences and allele frequency (**Supplementary Table 6**).^15^ A Firth logistic regression model was then applied for each model (with sex and age as covariates) to compare case carriers and control carriers. The significance threshold after Bonferroni correction for the number of genes and models tested in the 2 Mb region on chromosome 3 was α = (0.05 / [10 models×16 genes]) = 3.12×10^-4^; for the 1 Mb candidate region on chromosome 8 it was α = (0.05 / [10 models×10 genes]) = 5.00×10^-4^; and for the genome-wide exploration, it was α = (0.05 / [10 models×18,500genes]) = 2.70×10^-7^. Quantile-quantile plots and inflation factor (**Supplementary Figure 5**) were computed using the R package QQperm.^17^

### Functional variant annotation

We defined credible candidate variants as those located within 500 kb of the leading variants and with *P* values within two orders of magnitude of the lead variant. We used the software GCTA-CoJo^18^ to conduct a conditional analysis to identify independent signals. We used PAINTOR^19^ to identify credible sets of causal variants based on the magnitude and direction of the association and the pairwise linkage disequilibrium structure at the loci, and we used RegulomeDB^20^ and DSNetwork^21^ to assign a relative ranking to variants. We used *in silico* functional annotations from the public databases Open Target Genetics^22^, ExPheWAS^23^ and PhenoScanner v.2^24, 25^ to identify potential functional mechanisms and target genes. We tested the colocalization between the CHARM GWAS signals and clinically relevant phenotypes using the COLOC R package v3.2-1^26^ (refer to **Supplementary Material**).

### Data availability

The data underlying this article cannot be shared publicly to preserve the privacy of study participants. The analytic methods and study materials may be made available to other researchers for purposes of reproducing the results or replicating the procedure. Summary statistics from the GWAS analyses are available publicly for visualisation and download from **Supplementary Online Material** at https://pheweb.statgen.org/charm. The study protocol was approved by the Montreal Heart Institute research ethics committee and complies with the Declaration of Helsinki. Written informed consent was obtained from all participating subjects. The clinical trials upon which this post-hoc study is based had been registered on clinicaltrials.gov as NCT00634309, NCT00634712, and NCT00634400.

## RESULTS

There were 2727 participants available for the pharmacogenomic analysis of CHARM (**Figure 1**). The baseline characteristics of patients according to the study treatment groups are shown in **Table 1**. The mean age of participants was 66.5 years and 66.8 % were male. The primary endpoint of the individual CHARM studies, the composite of CV death or hospitalisation for HF, was used as the cardiovascular endpoint for the pharmacogenomic study, and occurred in 919 out of 2727 (33.7 %) participants in the pharmacogenomic sub-study, as compared to 2460 out of 7599 (32.4 %) patients in the overall CHARM programme.

**Table 1.**
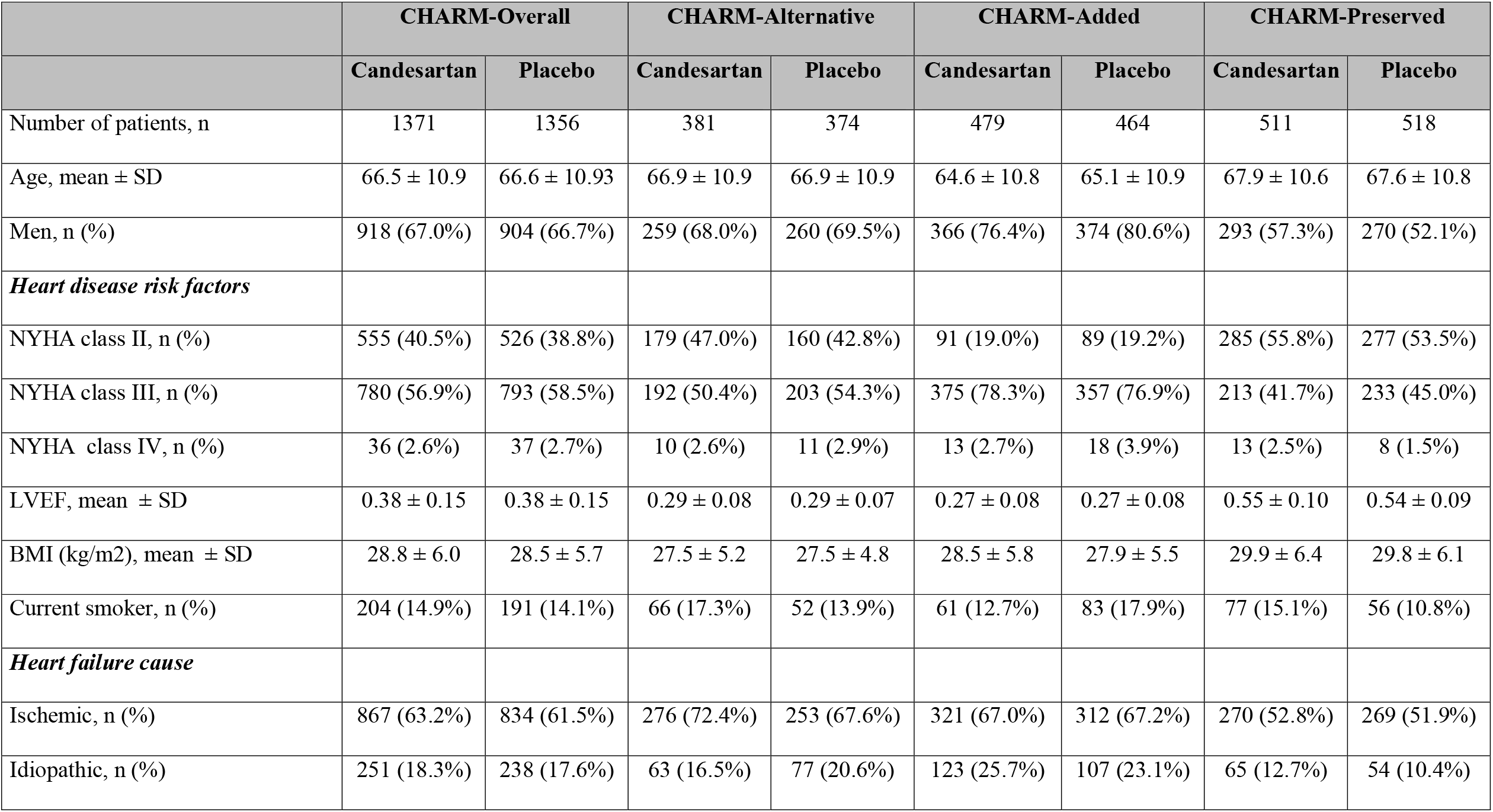
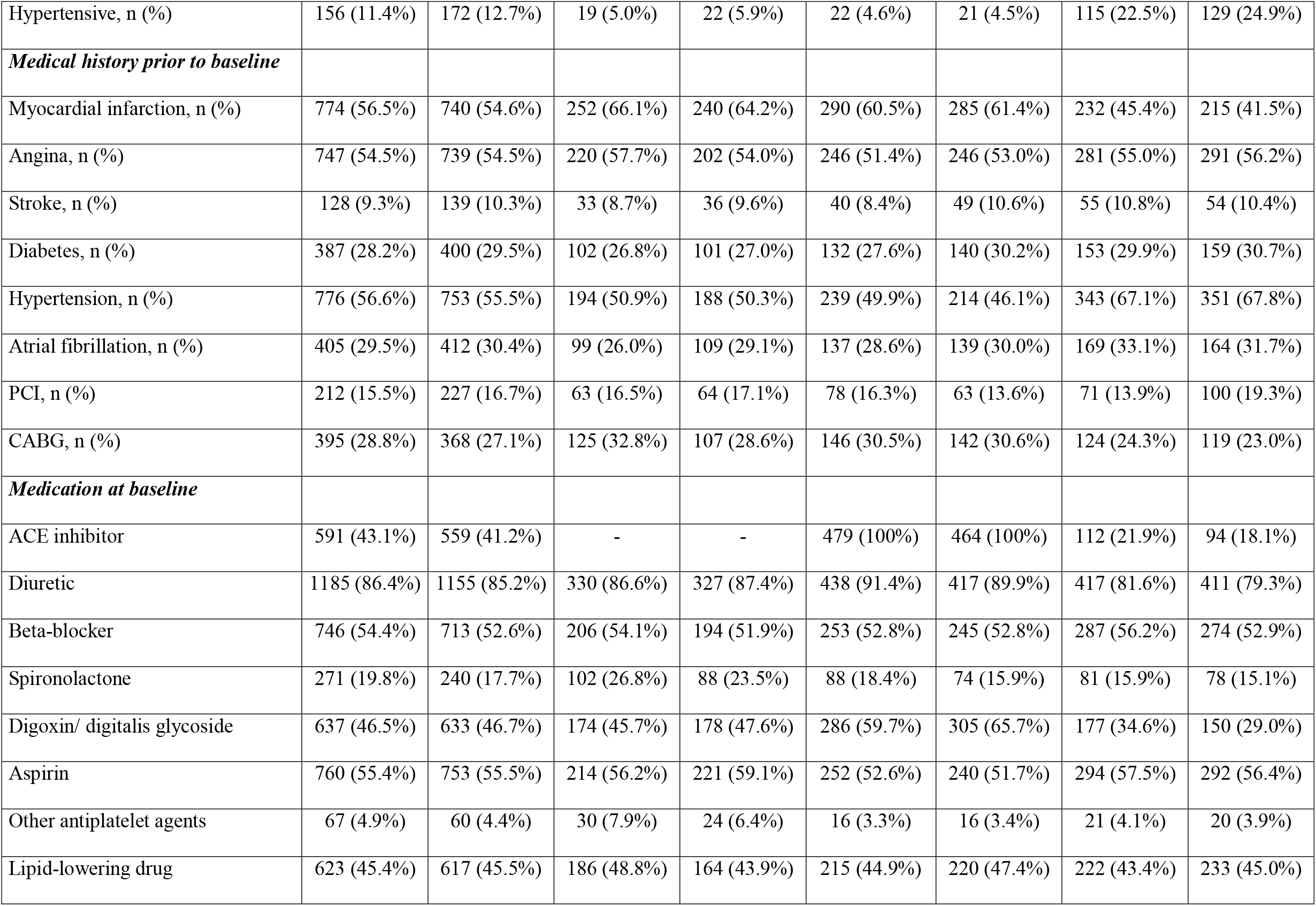
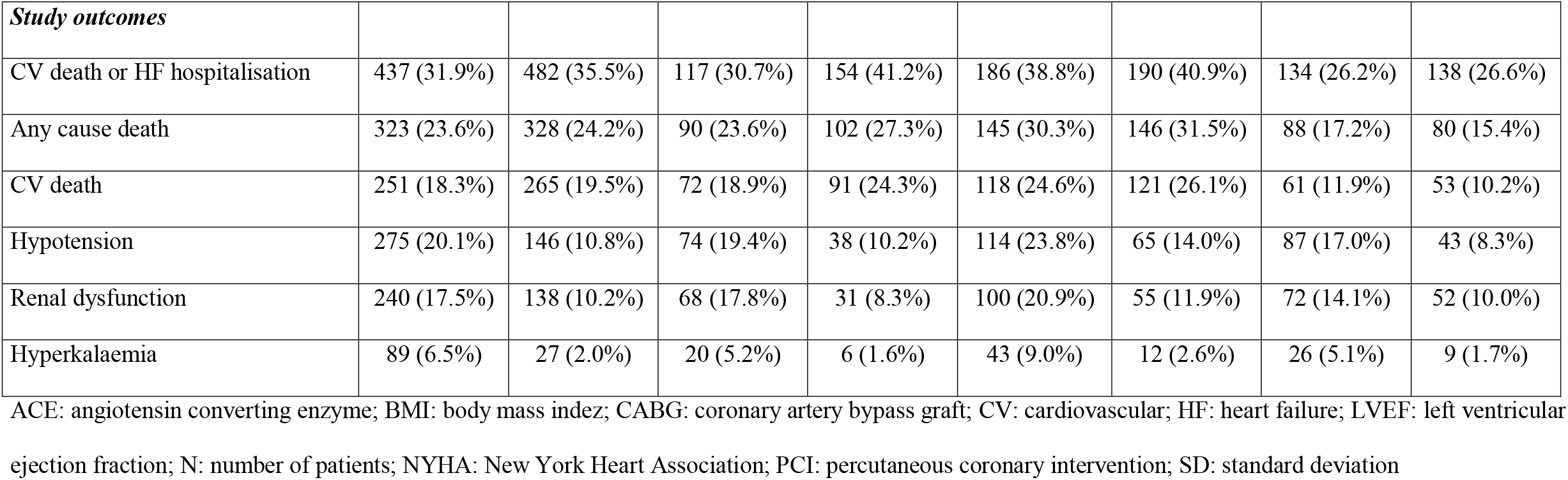
Characteristics of the pharmacogenomics study participants by CHARM study

|  | CHARM-Overall |  | CHARM-Alternative |  | CHARM-Added |  | CHARM-Preserved |  |
| --- | --- | --- | --- | --- | --- | --- | --- | --- |
|  | Candesartan | Placebo | Candesartan | Placebo | Candesartan | Placebo | Candesartan | Placebo |
| Number of patients, n | 1371 | 1356 | 381 | 374 | 479 | 464 | 511 | 518 |
| Age, mean ± SD | 66.5 ± 10.9 | 66.6 ± 10.93 | 66.9 ± 10.9 | 66.9 ± 10.9 | 64.6 ± 10.8 | 65.1 ± 10.9 | 67.9 ± 10.6 | 67.6 ± 10.8 |
| Men, n (%) | 918 (67.0%) | 904 (66.7%) | 259 (68.0%) | 260 (69.5%) | 366 (76.4%) | 374 (80.6%) | 293 (57.3%) | 270 (52.1%) |
| *Heart disease risk factors* |  |  |  |  |  |  |  |  |
| NYHA class II, n (%) | 555 (40.5%) | 526 (38.8%) | 179 (47.0%) | 160 (42.8%) | 91 (19.0%) | 89 (19.2%) | 285 (55.8%) | 277 (53.5%) |
| NYHA class III, n (%) | 780 (56.9%) | 793 (58.5%) | 192 (50.4%) | 203 (54.3%) | 375 (78.3%) | 357 (76.9%) | 213 (41.7%) | 233 (45.0%) |
| NYHA class IV, n (%) | 36 (2.6%) | 37 (2.7%) | 10 (2.6%) | 11 (2.9%) | 13 (2.7%) | 18 (3.9%) | 13 (2.5%) | 8 (1.5%) |
| LVEF, mean ± SD | 0.38 ± 0.15 | 0.38 ± 0.15 | 0.29 ± 0.08 | 0.29 ± 0.07 | 0.27 ± 0.08 | 0.27 ± 0.08 | 0.55 ± 0.10 | 0.54 ± 0.09 |
| BMI (kg/m<sup>2</sup>), mean ± SD | 28.8 ± 6.0 | 28.5 ± 5.7 | 27.5 ± 5.2 | 27.5 ± 4.8 | 28.5 ± 5.8 | 27.9 ± 5.5 | 29.9 ± 6.4 | 29.8 ± 6.1 |
| Current smoker, n (%) | 204 (14.9%) | 191 (14.1%) | 66 (17.3%) | 52 (13.9%) | 61 (12.7%) | 83 (17.9%) | 77 (15.1%) | 56 (10.8%) |
| *Heart failure cause* |  |  |  |  |  |  |  |  |
| Ischemic, n (%) | 867 (63.2%) | 834 (61.5%) | 276 (72.4%) | 253 (67.6%) | 321 (67.0%) | 312 (67.2%) | 270 (52.8%) | 269 (51.9%) |
| Idiopathic, n (%) | 251 (18.3%) | 238 (17.6%) | 63 (16.5%) | 77 (20.6%) | 123 (25.7%) | 107 (23.1%) | 65 (12.7%) | 54 (10.4%) |
| Hypertensive, n (%) | 156 (11.4%) | 172 (12.7%) | 19 (5.0%) | 22 (5.9%) | 22 (4.6%) | 21 (4.5%) | 115 (22.5%) | 129 (24.9%) |
| ***Medical history prior to baseline*** |  |  |  |  |  |  |  |  |
| Myocardial infarction, n (%) | 774 (56.5%) | 740 (54.6%) | 252 (66.1%) | 240 (64.2%) | 290 (60.5%) | 285 (61.4%) | 232 (45.4%) | 215 (41.5%) |
| Angina, n (%) | 747 (54.5%) | 739 (54.5%) | 220 (57.7%) | 202 (54.0%) | 246 (51.4%) | 246 (53.0%) | 281 (55.0%) | 291 (56.2%) |
| Stroke, n (%) | 128 (9.3%) | 139 (10.3%) | 33 (8.7%) | 36 (9.6%) | 40 (8.4%) | 49 (10.6%) | 55 (10.8%) | 54 (10.4%) |
| Diabetes, n (%) | 387 (28.2%) | 400 (29.5%) | 102 (26.8%) | 101 (27.0%) | 132 (27.6%) | 140 (30.2%) | 153 (29.9%) | 159 (30.7%) |
| Hypertension, n (%) | 776 (56.6%) | 753 (55.5%) | 194 (50.9%) | 188 (50.3%) | 239 (49.9%) | 214 (46.1%) | 343 (67.1%) | 351 (67.8%) |
| Atrial fibrillation, n (%) | 405 (29.5%) | 412 (30.4%) | 99 (26.0%) | 109 (29.1%) | 137 (28.6%) | 139 (30.0%) | 169 (33.1%) | 164 (31.7%) |
| PCI, n (%) | 212 (15.5%) | 227 (16.7%) | 63 (16.5%) | 64 (17.1%) | 78 (16.3%) | 63 (13.6%) | 71 (13.9%) | 100 (19.3%) |
| CABG, n (%) | 395 (28.8%) | 368 (27.1%) | 125 (32.8%) | 107 (28.6%) | 146 (30.5%) | 142 (30.6%) | 124 (24.3%) | 119 (23.0%) |
| ***Medication at baseline*** |  |  |  |  |  |  |  |  |
| ACE inhibitor | 591 (43.1%) | 559 (41.2%) | - | - | 479 (100%) | 464 (100%) | 112 (21.9%) | 94 (18.1%) |
| Diuretic | 1185 (86.4%) | 1155 (85.2%) | 330 (86.6%) | 327 (87.4%) | 438 (91.4%) | 417 (89.9%) | 417 (81.6%) | 411 (79.3%) |
| Beta-blocker | 746 (54.4%) | 713 (52.6%) | 206 (54.1%) | 194 (51.9%) | 253 (52.8%) | 245 (52.8%) | 287 (56.2%) | 274 (52.9%) |
| Spironolactone | 271 (19.8%) | 240 (17.7%) | 102 (26.8%) | 88 (23.5%) | 88 (18.4%) | 74 (15.9%) | 81 (15.9%) | 78 (15.1%) |
| Digoxin/ digitalis glycoside | 637 (46.5%) | 633 (46.7%) | 174 (45.7%) | 178 (47.6%) | 286 (59.7%) | 305 (65.7%) | 177 (34.6%) | 150 (29.0%) |
| Aspirin | 760 (55.4%) | 753 (55.5%) | 214 (56.2%) | 221 (59.1%) | 252 (52.6%) | 240 (51.7%) | 294 (57.5%) | 292 (56.4%) |
| Other antiplatelet agents | 67 (4.9%) | 60 (4.4%) | 30 (7.9%) | 24 (6.4%) | 16 (3.3%) | 16 (3.4%) | 21 (4.1%) | 20 (3.9%) |
| Lipid-lowering drug | 623 (45.4%) | 617 (45.5%) | 186 (48.8%) | 164 (43.9%) | 215 (44.9%) | 220 (47.4%) | 222 (43.4%) | 233 (45.0%) |
| *Study outcomes* |  |  |  |  |  |  |  |  |
| CV death or HF hospitalisation | 437 (31.9%) | 482 (35.5%) | 117 (30.7%) | 154 (41.2%) | 186 (38.8%) | 190 (40.9%) | 134 (26.2%) | 138 (26.6%) |
| Any cause death | 323 (23.6%) | 328 (24.2%) | 90 (23.6%) | 102 (27.3%) | 145 (30.3%) | 146 (31.5%) | 88 (17.2%) | 80 (15.4%) |
| CV death | 251 (18.3%) | 265 (19.5%) | 72 (18.9%) | 91 (24.3%) | 118 (24.6%) | 121 (26.1%) | 61 (11.9%) | 53 (10.2%) |
| Hypotension | 275 (20.1%) | 146 (10.8%) | 74 (19.4%) | 38 (10.2%) | 114 (23.8%) | 65 (14.0%) | 87 (17.0%) | 43 (8.3%) |
| Renal dysfunction | 240 (17.5%) | 138 (10.2%) | 68 (17.8%) | 31 (8.3%) | 100 (20.9%) | 55 (11.9%) | 72 (14.1%) | 52 (10.0%) |
| Hyperkalaemia | 89 (6.5%) | 27 (2.0%) | 20 (5.2%) | 6 (1.6%) | 43 (9.0%) | 12 (2.6%) | 26 (5.1%) | 9 (1.7%) |

ACE: angiotensin converting enzyme; BMI: body mass index; CABG: coronary artery bypass graft; CV: cardiovascular; HF: heart failure; LVEF: left ventricular ejection fraction; N: number of patients; NYHA: New York Heart Association; PCI: percutaneous coronary intervention; SD: standard deviation

**Table 2.** Significant GWAS association results for the composite of cardiovascular death or hospitalisation for heart failure in the CHARM-Preserved study

| Genetic variant | Chr | Position | Nearest coding gene | EA | RA | EAF | N | N events | HR* (95% CI) | P value |
| --- | --- | --- | --- | --- | --- | --- | --- | --- | --- | --- |
| rs112455636 | 8 | 21,599,237 | *GFRA2* | T | C | 0.141 | 1,019 | 269 | 1.91 (1.54, 2.37) | $2.7 \times 10^{-9}$ |
| rs144872887 | 8 | 21,605,938 | *GFRA2* | C | CAG | 0.144 | 1,024 | 270 | 1.90 (1.54, 2.35) | $1.8 \times 10^{-9}$ |
| **rs66886237** | **8** | **21,607,231** | ***GFRA2*** | **A** | **G** | **0.144** | **1,024** | **270** | **1.91 (1.55, 2.35)** | **$1.7 \times 10^{-9}$** |

Position on build37. Chr: chromosome; CI: confidence interval; EA: effect allele; EAF: effect allele frequency in the analysis dataset; HF: heart failure; RA: reference allele; HR: hazard ratio.
\* Allelic dosage effect assessed using Cox proportional hazards regression adjusted for age, sex, 10 principal components and candesartan treatment.

### GWAS of progression of CV endpoints in HF

We conducted 3 GWAS to identify genetic variants associated with the composite of CV death or hospitalisation for HF as an indicator of disease progression using 1) patients from the CHARM-Overall programme, 2) the subset of patients with reduced ejection fraction from the CHARM-Alternative and CHARM-Added studies, and 3) the subset of patients with preserved ejection fraction from the CHARM-preserved study. We found a significant association signal in the GWAS of HF patients with preserved ejection fraction (**Figure 2**). There were 1029 patients included in the analysis, of which 272 (26.4%) had an event. The genetic variant rs66886237 at 8p21.3 is leading the signal with HR=1.91 (95% IC: 1.55-2.35), P=1.7×10^-9^. When conditioning on rs66886237, no additional genetic variants remained significant at P<2.8×10^-9^ in the region, and rs66886237 ranked as the best functional candidate in the region (**Supplementary material**). The genetic variant was not associated with the composite of CV death or hospitalisation for HF in patients with reduced ejection fraction from the CHARM-Alternative and CHARM-Added studies (P>0.05; **Table 3**). The effect of rs66886237 was not modulated by sex or by treatment with candesartan. Sensitivity analyses were conducted to assess the association of rs66886237 with the composite of CV death or hospitalisation for HF in CHARM-Preserved with further adjustment (individually) for BMI, left ventricular ejection fraction, history of atrial fibrillation, and prior MI, diabetes, and in all of the tested models, the genetic variant remained significant at P<2.8×10^-9^. Baseline characteristics and study outcomes are presented according to rs66886237 genotypes in **Supplementary Table 3**. The cumulative incidence of the composite of CV death or hospitalisation for heart failure is presented in **Figure 3** stratified according to rs66886237 genotypes. The minor allele (A) had a frequency of 14% in the study population. The variant is located in an intron of the *GFRA2* gene encoding the glial cell line-derived neurotrophic factor (GDNF) family receptor alpha 2.

**Figure 2.**
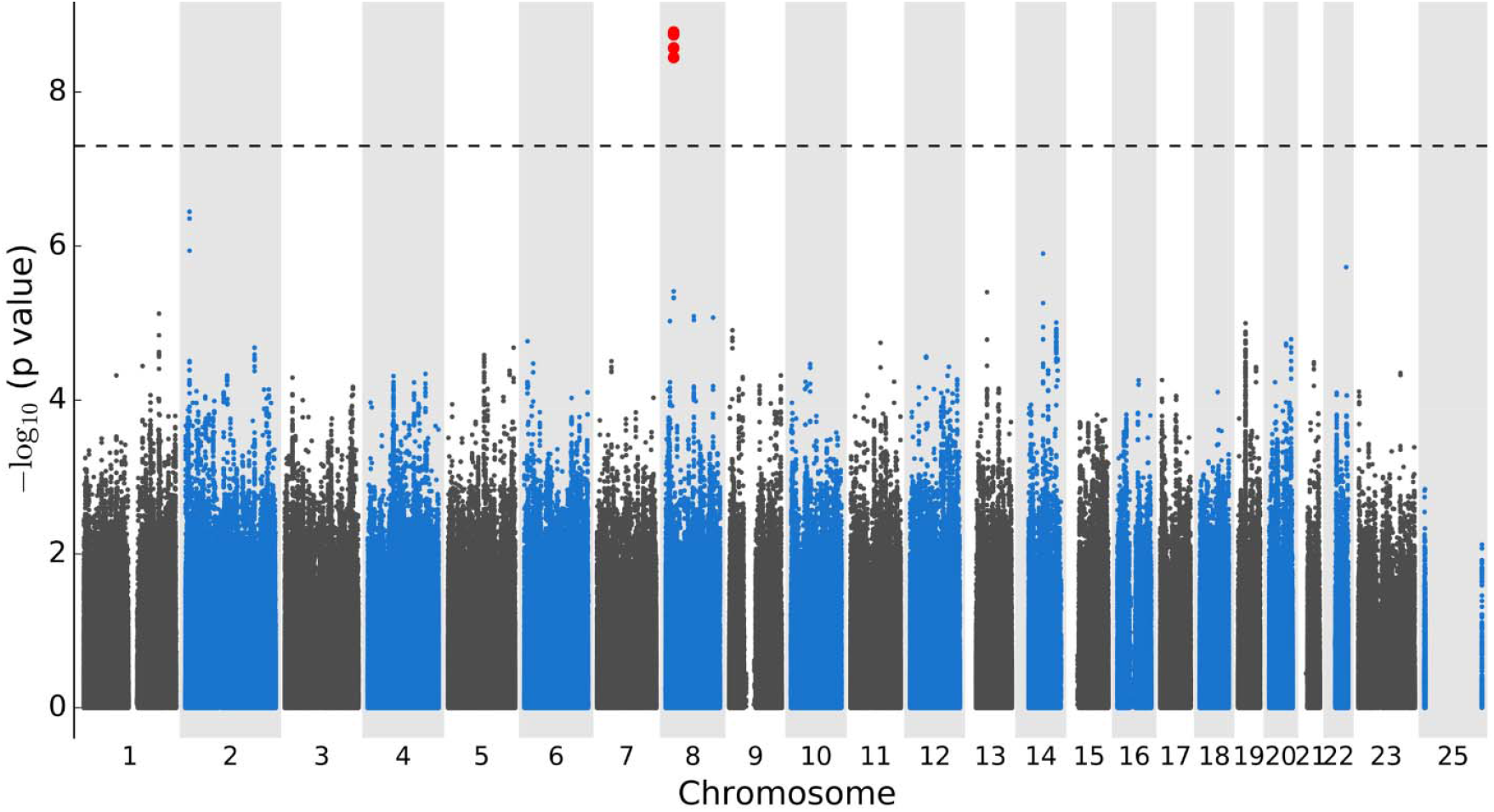
Manhattan plot of the GWAS with 1029 heart failure patients with preserved ejection fraction from the CHARM-Preserved study tested for association with time to cardiovascular death or heart failure hospitalisation using Cox proportional hazards regression with adjustment for principal components for genetic ancestry, treatment arm, age, and sex. There were 5,023,375 genotyped and imputed genetic variants of MAF ≥ 5%. The dashed line marks P=5×10^-8^.

**Table 3.** Association results for the composite of cardiovascular death or heart failure hospitalisation in the CHARM studies for leading genetic variant rs66886237 at 8p21.3

| Genetic variant<br>:effect allele | CHARM studies | N | N Events (%) | HR* (95% CI) | P value |
| --- | --- | --- | --- | --- | --- |
| rs66886237:A | Overall | 2713 | 912 (33.62%) | 1.17 (1.02-1.33) | 0.0237 |
|  | Alternative | 754 | 270 (35.81%) | 0.92 (0.70-1.20) | 0.5182 |
|  | Added | 935 | 372 (39.79%) | 0.83 (0.66-1.05) | 0.1294 |
|  | Alternative + Added | 1689 | 642 (38.01%) | 0.88 (0.74-1.05) | 0.1629 |
| | Preserved | 1024 | 270 (26.37%) | 1.91 (1.55-2.35) | $1.7 \times 10^{-9}$ |

CI: confidence interval; HR: hazard ratio; N: number of patients
\* Allelic dosage effect assessed using Cox proportional hazards regression adjusted for age, sex, 10 principal components and candesartan treatment.

**Figure 3.**
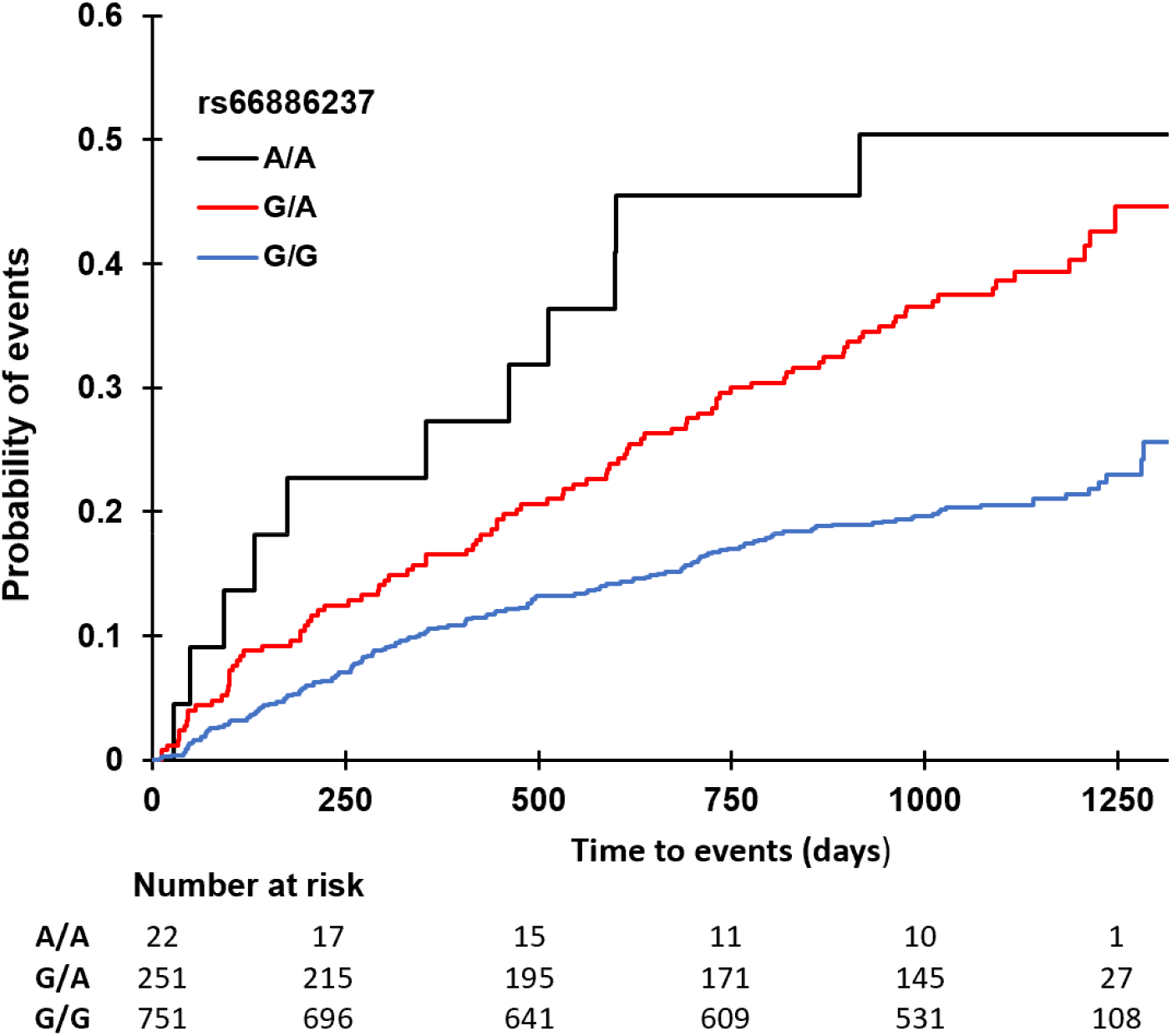
Cumulative incidence curves for the composite endpoint of cardiovascular death or heart failure hospitalisation in CHARM-Preserved study participants stratified by rs66886237 genotypes.

### GWAS of candesartan efficacy on CV endpoints

We conducted 3 GWAS to discover genetic variants predictive of candesartan efficacity with the composite CV endpoint using the candesartan arm of the CHARM-Overall programme, the combined CHARM-Alternative and CHARM-Added studies, and the CHARM-Preserved study. No results passed the multiple-testing corrected GWAS significance threshold (P<2.8×10^-9^). One region on chromosome 3 at 3q13.13 passed the unadjusted threshold of P<5×10^-8^, with variant rs664669 leading the signal in the CHARM-Overall programme in 1371 participants randomised to candesartan (**Supplementary Figure 2, Supplementary Table 4**). The minor allele (C) had a frequency of 44% and was associated with the composite endpoint of CV death or hospitalisation for HF in the candesartan group (HR= 1.48, 95% CI 1.29-1.69, P= 2.63×10^-8^) with no effect in the placebo group (HR= 1.11, 95% CI 0.97-1.26; P= 0.125) and the interaction term between rs664669 and candesartan treatment was significant (P= 0.003) (**Supplementary Table 4**). Variant rs664669 is intronic to the long interspersed non-coding RNA *RP11-457K10*.*1*, a processed pseudo gene of 763 base pairs with a transcript of 262 base pairs. Exploratory GWAS testing for the interaction effect between genome-wide variants and treatment also did not find significant signals (**Supplementary Online Material**).

### GWAS of candesartan safety endpoints

We conducted 15 GWAS to identify genetic variants predictive of hyperkalaemia, renal dysfunction, hypotension, and change in systolic blood pressure in candesartan-treated patients. There were 1371 participants randomised to candesartan in all three CHARM studies who were included in the genetic analyses of the GWAS with safety endpoints, of those, 89 (6.5 %) patients had a report of hyperkalaemia adverse events, 240 (17.5 %) with renal dysfunction, and 275 (20.1 %) with hypotension. None of the tested genetic variants passed the GWAS significance threshold (P<2.8×10^-9^). Exploratory GWAS testing for the interaction effect between genome-wide variants and treatment also did not reveal significant signals (**Supplementary Online Material**).

### Whole-exome sequencing data association with the CV endpoint

We conducted a genome-wide gene-level collapsing analysis for the composite of CV death or hospitalisation for HF in the CHARM-Preserved study and in the CHARM-Overall study to support the GWAS findings. None of the 18,500 tested coding genes was significantly associated with the composite cardiovascular endpoint in candesartan treated patients (**Supplementary Table 7**). In the candidate regions at 8p21.3 and 3q13.13, none of the genes within the flanking regions were significantly associated (**Supplementary Table 8**).

### Candidate genetic variant AGTR1 rs5186

Multiple genetic variants related to the renin-angiotensin system have been proposed to modulate the effects of ARBs in HF and other cardiovascular diseases. The *AGTR1* rs5186 genetic variant, which results in an A to C substitution at position 1166 in the 3’-untranslated region of the *AGTR1* gene, has arguably been to the most widely studied. This variant corresponds to the binding site for miR155 on the messenger RNA, which ultimately produces an RNA-silencing complex and an inhibition of translation in the presence of the A1166 allele. The C1166 allele is expected to be associated with the highest expression of the receptor. We found no significant association, even at a nominal level, with the composite cardiovascular endpoint in the overall CHARM programme or combined CHARM-Alternative and CHARM-Added analyses, nor with the risk of hyperkalaemia, renal dysfunction or changes in SBP. The only nominal association observed was with the risk of hypotension in the combined CHARM-Alternative and CHARM-Added analyses (P= 0.029).

## DISCUSSION

In this pharmacogenomic study of the CHARM programme, we have identified a candidate genetic predictor of HF progression in patients with preserved ejection fraction from the CHARM-Preserved study with the composite of CV death or hospitalisation for HF. The genetic variant rs66886237-A at 8p21.3 was associated with time to hospitalisation for HF or CV death with an allelic HR of 1.91. The lead variant at this locus is located in an intron of the *GFRA2* gene. Functional analysis of the variant did not find it to be a regulator of gene expression and it was not previously strongly associated with phenotypes in queried databases. However, the genetic region identified was concordant by colocalization analysis with a locus associated with cardiomyopathy in the FinnGen project, based on hospital discharge and cause of death registries (ICD-10 code I42).^27^ *Gfra2* was found to be a specific marker for cardiac progenitors among mesodermal cells and mice, and *GFRA2* expression marks human developmental cardiac progenitor cell populations in embryonic stem cells/induced pluripotent stem cells differentiation.^28^

We also report a non-significant association signal for the efficacy of candesartan at 3q13.13. Individuals with the rs664669-TT genotype represented approximately 31% of the population and they had a 41% reduction (95% CI: 0.45-0.76) in the combined incidence of CV death and hospitalisation for HF when treated with candesartan as compared to placebo. This may represent further benefit to that observed in the overall population with 16% reduction in CV death and hospitalisation for HF reported in the CHARM-Overall programme. However, the finding is hypothesis-generating, and replication of the association in an independent population sample is necessary.

The previously reported candidate gene for response to candesartan, the angiotensin II receptor type 1 gene (*AGTR1)*,^29, 30^ was not associated with the tested endpoints, except for a possible association with hypotension in the HFrEF patient population. Based on our data, it is unlikely that single common genetic variants will have a major impact on response to candesartan. Nonetheless, given the limited size of the study population and the complexity of the HF phenotype, we cannot exclude the existence of a modest effect of several other gene variants including that of *AGTR1*.

Our study had limitations. There may be volunteer bias in the pharmacogenomic subgroup compared to the main trial population, but the subgroup of participants was representative of the main trial in terms of the composite cardiovascular endpoint of time to CV death or hospitalisation for HF. Although we have corrected for the testing of several phenotypes, the results are to be interpreted with care and should be considered strictly as hypothesis generating. Results will have to be replicated and demonstrate biological plausibility before considering any changes in clinical practice. We have also limited our analyses to study subjects genetically predicted to be of European ancestry by comparison to HapMap CEU samples. This was done to protect from population structure which may bias results. We did not have enough study participants from other population groups to assess the generalisability of our finding across other ancestries. The leading variant rs66886237 had a minor allele frequency of 0.14 in our study population, which compares to the frequency of 0.15 in samples from European populations in the 1000 Genomes reference,^31^ and minor allele frequencies of 0.41 and 0.09 in samples from African and East Asian populations respectively. Importantly, the results have not been replicated in an independent population sample, and we cannot exclude the possibility that the results may be chance findings.

In conclusion, we have identified a candidate genetic region at 8p21.3 near gene *GFRA2* associated with the progression of HF in patients with preserved ejection fraction from the CHARM-Preserved study. The findings will need to be replicated in an independent population.

## Data Availability

The data underlying this article cannot be shared publicly to preserve the privacy of study participants. The analytic methods and study materials may be made available to other researchers for purposes of reproducing the results or replicating the procedure. Summary statistics from the GWAS analyses are available publicly for visualisation and download at https://pheweb.statgen.org/charm

## ACKNOWLEDGMENTS

We thank the patients and all clinical and research personnel who enable the conduct of the CHARM studies. The sequencing analysis was performed in the Centre for Genomics Research (CGR), Discovery Sciences, BioPharmaceuticals R&D, AstraZeneca, Cambridge, UK. We would like to thank Slavé Petrovski and Sri Deevi from the CGR for their support, for which we are grateful.

## REFERENCES

1. McKelvie RS, Yusuf S, Pericak D, Avezum A, Burns RJ, Probstfield J, Tsuyuki RT, White M, Rouleau J, Latini R, Maggioni A, Young J, Pogue J. Comparison of candesartan, enalapril, and their combination in congestive heart failure: randomized evaluation of strategies for left ventricular dysfunction (RESOLVD) pilot study. The RESOLVD Pilot Study Investigators. Circulation 1999;100(10):1056–64.

2. Tsutamoto T, Wada A, Maeda K, Mabuchi N, Hayashi M, Tsutsui T, Ohnishi M, Sawaki M, Fujii M, Matsumoto T, Kinoshita M. Angiotensin II type 1 receptor antagonist decreases plasma levels of tumor necrosis factor alpha, interleukin-6 and soluble adhesion molecules in patients with chronic heart failure. J Am Coll Cardiol 2000;35(3):714–21.

3. White M, Lepage S, Lavoie J, De Denus S, Leblanc MH, Gossard D, Whittom L, Racine N, Ducharme A, Dabouz F, Rouleau JL, Touyz R. Effects of combined candesartan and ACE inhibitors on BNP, markers of inflammation and oxidative stress, and glucose regulation in patients with symptomatic heart failure. J Card Fail 2007;13(2):86–94.

4. Granger CB, McMurray JJ, Yusuf S, Held P, Michelson EL, Olofsson B, Ostergren J, Pfeffer MA, Swedberg K, Investigators C, Committees. Effects of candesartan in patients with chronic heart failure and reduced left-ventricular systolic function intolerant to angiotensin-converting-enzyme inhibitors: the CHARM-Alternative trial. Lancet 2003;362(9386):772–6.

5. McMurray JJ, Ostergren J, Swedberg K, Granger CB, Held P, Michelson EL, Olofsson B, Yusuf S, Pfeffer MA, Investigators C, Committees. Effects of candesartan in patients with chronic heart failure and reduced left-ventricular systolic function taking angiotensin-converting-enzyme inhibitors: the CHARM-Added trial. Lancet 2003;362(9386):767–71.

6. Yusuf S, Pfeffer MA, Swedberg K, Granger CB, Held P, McMurray JJ, Michelson EL, Olofsson B, Ostergren J, Investigators C, Committees. Effects of candesartan in patients with chronic heart failure and preserved left-ventricular ejection fraction: the CHARM-Preserved Trial. Lancet 2003;362(9386):777–81.

7. Cohn JN, Tognoni G. A randomized trial of the angiotensin-receptor blocker valsartan in chronic heart failure. N Engl J Med 2001;345(23):1667–75.

8. Swedberg K, Pfeffer M, Granger C, Held P, McMurray J, Ohlin G, Olofsson B, Ostergren J, Yusuf S. Candesartan in heart failure--assessment of reduction in mortality and morbidity (CHARM): rationale and design. Charm-Programme Investigators. J Card Fail 1999;5(3):276–82.

9. Pfeffer MA, Swedberg K, Granger CB, Held P, McMurray JJ, Michelson EL, Olofsson B, Ostergren J, Yusuf S, Pocock S, Investigators C, Committees. Effects of candesartan on mortality and morbidity in patients with chronic heart failure: the CHARM-Overall programme. Lancet 2003;362(9386):759–66.

10. Lemieux Perreault LP, Provost S, Legault MA, Barhdadi A, Dube MP. pyGenClean: efficient tool for genetic data clean up before association testing. Bioinformatics 2013;29(13):1704–5.

11. Purcell S, Neale B, Todd-Brown K, Thomas L, Ferreira MA, Bender D, Maller J, Sklar P, de Bakker PI, Daly MJ, Sham PC. PLINK: a tool set for whole-genome association and population-based linkage analyses. Am J Hum Genet 2007;81(3):559–75.

12. Price AL, Patterson NJ, Plenge RM, Weinblatt ME, Shadick NA, Reich D. Principal components analysis corrects for stratification in genome-wide association studies. Nat Genet 2006;38(8):904–9.

13. Howie B, Marchini J, Stephens M. Genotype imputation with thousands of genomes. G3 (Bethesda) 2011;1(6):457–70.

14. Delaneau O, Marchini J, Zagury JF. A linear complexity phasing method for thousands of genomes. Nat Methods 2012;9(2):179–81.

15. Carss KJ, Baranowska AA, Armisen J, Webb TR, Hamby SE, Premawardhana D, Al-Hussaini A, Wood A, Wang Q, Deevi SVV, Vitsios D, Lewis SH, Kotecha D, Bouatia-Naji N, Hesselson S, Iismaa SE, Tarr I, McGrath-Cadell L, Muller DW, Dunwoodie SL, Fatkin D, Graham RM, Giannoulatou E, Samani NJ, Petrovski S, Haefliger C, Adlam D. Spontaneous Coronary Artery Dissection: Insights on Rare Genetic Variation From Genome Sequencing. Circ Genom Precis Med 2020;13(6):e003030.

16. Carss KJ, Baranowska AA, Armisen J, Webb TR, Hamby SE, Premawardhana D, Al-Hussaini A, Wood A, Wang Q, Deevi SVV, Vitsios D, Lewis SH, Kotecha D, Bouatia-Naji N, Hesselson S, Iismaa SE, Tarr I, McGrath-Cadell L, Muller DW, Dunwoodie SL, Fatkin D, Graham RM, Giannoulatou E, Samani NJ, Petrovski S, Haefliger C, Adlam D. Spontaneous Coronary Artery Dissection: Insights on Rare Genetic Variation from Genome Sequencing. Circ Genom Precis Med 2020.

17. Slave Petrovski QW. QQperm: permutation based QQ plot and inflation factor estimation. https://cran.r-project.org/web/packages/QQperm/index.html.

18. Yang J, Lee SH, Goddard ME, Visscher PM. GCTA: a tool for genome-wide complex trait analysis. Am J Hum Genet 2011;88(1):76–82.

19. Kichaev G, Roytman M, Johnson R, Eskin E, Lindstrom S, Kraft P, Pasaniuc B. Improved methods for multi-trait fine mapping of pleiotropic risk loci. Bioinformatics 2017;33(2):248–255.

20. Boyle AP, Hong EL, Hariharan M, Cheng Y, Schaub MA, Kasowski M, Karczewski KJ, Park J, Hitz BC, Weng S, Cherry JM, Snyder M. Annotation of functional variation in personal genomes using RegulomeDB. Genome Res 2012;22(9):1790–7.

21. Lemacon A, Scott-Boyer MP, Ongaro-Carcy R, Soucy P, Simard J, Droit A. DSNetwork: An Integrative Approach to Visualize Predictions of Variants’ Deleteriousness. Front Genet 2019;10:1349.

22. Ghoussaini M, Mountjoy E, Carmona M, Peat G, Schmidt EM, Hercules A, Fumis L, Miranda A, Carvalho-Silva D, Buniello A, Burdett T, Hayhurst J, Baker J, Ferrer J, Gonzalez-Uriarte A, Jupp S, Karim MA, Koscielny G, Machlitt-Northen S, Malangone C, Pendlington ZM, Roncaglia P, Suveges D, Wright D, Vrousgou O, Papa E, Parkinson H, MacArthur JAL, Todd JA, Barrett JC, Schwartzentruber J, Hulcoop DG, Ochoa D, McDonagh EM, Dunham I. Open Targets Genetics: systematic identification of trait-associated genes using large-scale genetics and functional genomics. Nucleic Acids Res 2021;49(D1):D1311–D1320.

23. Legault M-A, Perreault L-PL, Dubé M-P. ExPheWas: a browser for gene-based pheWAS associations. medRxiv 2021:2021.03.17.21253824.

24. Kamat MA, Blackshaw JA, Young R, Surendran P, Burgess S, Danesh J, Butterworth AS, Staley JR. PhenoScanner V2: an expanded tool for searching human genotype-phenotype associations. Bioinformatics 2019;35(22):4851–4853.

25. Staley JR, Blackshaw J, Kamat MA, Ellis S, Surendran P, Sun BB, Paul DS, Freitag D, Burgess S, Danesh J, Young R, Butterworth AS. PhenoScanner: a database of human genotype-phenotype associations. Bioinformatics 2016;32(20):3207–3209.

26. Wallace C. Eliciting priors and relaxing the single causal variant assumption in colocalisation analyses. PLoS Genet 2020;16(4):e1008720.

27. FinnGen. Documentation of R4 release. https://finngen.gitbook.io/documentation/.

28. Ishida H, Saba R, Kokkinopoulos I, Hashimoto M, Yamaguchi O, Nowotschin S, Shiraishi M, Ruchaya P, Miller D, Harmer S, Poliandri A, Kogaki S, Sakata Y, Dunkel L, Tinker A, Hadjantonakis A-K, Sawa Y, Sasaki H, Ozono K, Suzuki K, Yashiro K. GFRA2 Identifies Cardiac Progenitors and Mediates Cardiomyocyte Differentiation in a RET-Independent Signaling Pathway. Cell Reports 2016;16(4):1026–1038.

29. de Denus S, Zakrzewski-Jakubiak M, Dube MP, Belanger F, Lepage S, Leblanc MH, Gossard D, Ducharme A, Racine N, Whittom L, Lavoie J, Touyz RM, Turgeon J, White M. Effects of AGTR1 A1166C gene polymorphism in patients with heart failure treated with candesartan. Ann Pharmacother 2008;42(7):925–32.

30. de Denus S, Dube MP, Fouodjio R, Huynh T, LeBlanc MH, Lepage S, Sheppard R, Giannetti N, Lavoie J, Mansour A, Provost S, Normand V, Mongrain I, Langlois M, O’Meara E, Ducharme A, Racine N, Guertin MC, Turgeon J, Phillips MS, Rouleau JL, Tardif JC, White M, investigators CI. A prospective study of the impact of AGTR1 A1166C on the effects of candesartan in patients with heart failure. Pharmacogenomics 2018;19(7):599–612.

31. Auton A, Abecasis GR, Altshuler DM, Durbin RM, Abecasis GR, Bentley DR, Chakravarti A, Clark AG, Donnelly P, Eichler EE, Flicek P, Gabriel SB, Gibbs RA, Green ED, Hurles ME, Knoppers BM, Korbel JO, Lander ES, Lee C, Lehrach H, Mardis ER, Marth GT, McVean GA, Nickerson DA, Schmidt JP, Sherry ST, Wang J, Wilson RK, Gibbs RA, Boerwinkle E, Doddapaneni H, Han Y, Korchina V, Kovar C, Lee S, Muzny D, Reid JG, Zhu Y, Wang J, Chang Y, Feng Q, Fang X, Guo X, Jian M, Jiang H, Jin X, Lan T, Li G, Li J, Li Y, Liu S, Liu X, Lu Y, Ma X, Tang M, Wang B, Wang G, Wu H, Wu R, Xu X, Yin Y, Zhang D, Zhang W, Zhao J, Zhao M, Zheng X, Lander ES, Altshuler DM, Gabriel SB, Gupta N, Gharani N, Toji LH, Gerry NP, Resch AM, Flicek P, Barker J, Clarke L, Gil L, Hunt SE, Kelman G, Kulesha E, Leinonen R, McLaren WM, Radhakrishnan R, Roa A, Smirnov D, Smith RE, Streeter I, Thormann A, Toneva I, Vaughan B, Zheng-Bradley X, Bentley DR, Grocock R, Humphray S, James T, Kingsbury Z, Lehrach H, Sudbrak R, Albrecht MW, Amstislavskiy VS, Borodina TA, Lienhard M, Mertes F, Sultan M, Timmermann B, Yaspo M-L, Mardis ER, Wilson RK, Fulton L, Fulton R, Sherry ST, Ananiev V, Belaia Z, Beloslyudtsev D, Bouk N, Chen C, Church D, Cohen R, Cook C, Garner J, Hefferon T, Kimelman M, Liu C, Lopez J, Meric P, O’Sullivan C, Ostapchuk Y, Phan L, Ponomarov S, Schneider V, Shekhtman E, Sirotkin K, Slotta D, Zhang H, McVean GA, Durbin RM, Balasubramaniam S, Burton J, Danecek P, Keane TM, Kolb-Kokocinski A, McCarthy S, Stalker J, Quail M, Schmidt JP, Davies CJ, Gollub J, Webster T, Wong B, Zhan Y, Auton A, Campbell CL, Kong Y, Marcketta A, Gibbs RA, Yu F, Antunes L, Bainbridge M, Muzny D, Sabo A, Huang Z, Wang J, Coin LJM, Fang L, Guo X, Jin X, Li G, Li Q, Li Y, Li Z, Lin H, Liu B, Luo R, Shao H, Xie Y, Ye C, Yu C, Zhang F, Zheng H, Zhu H, Alkan C, Dal E, Kahveci F, Marth GT, Garrison EP, Kural D, Lee W-P, Fung Leong W, Stromberg M, Ward AN, Wu J, Zhang M, Daly MJ, DePristo MA, Handsaker RE, Altshuler DM, Banks E, Bhatia G, del Angel G, Gabriel SB, Genovese G, Gupta N, Li H, Kashin S, Lander ES, McCarroll SA, Nemesh JC, Poplin RE, Yoon SC, Lihm J, Makarov V, Clark AG, Gottipati S, Keinan A, Rodriguez-Flores JL, Korbel JO, Rausch T, Fritz MH, Stütz AM, Flicek P, Beal K, Clarke L, Datta A, Herrero J, McLaren WM, Ritchie GRS, Smith RE, Zerbino D, Zheng-Bradley X, Sabeti PC, Shlyakhter I, Schaffner SF, Vitti J, Cooper DN, Ball EV, Stenson PD, Bentley DR, Barnes B, Bauer M, Keira Cheetham R, Cox A, Eberle M, Humphray S, Kahn S, Murray L, Peden J, Shaw R, Kenny EE, Batzer MA, Konkel MK, Walker JA, MacArthur DG, Lek M, Sudbrak R, Amstislavskiy VS, Herwig R, Mardis ER, Ding L, Koboldt DC, Larson D, Ye K, Gravel S, The Genomes Project C, Corresponding a, Steering c, Production g, Baylor College of M, Shenzhen BGI, Broad Institute of MIT, Harvard, Coriell Institute for Medical R, European Molecular Biology Laboratory EBI, Illumina, Max Planck Institute for Molecular G, McDonnell Genome Institute at Washington U, Health USNIo, University of O, Wellcome Trust Sanger I, Analysis g, Affymetrix, Albert Einstein College of M, Bilkent U, Boston C, Cold Spring Harbor L, Cornell U, European Molecular Biology L, Harvard U, Human Gene Mutation D, Icahn School of Medicine at Mount S, Louisiana State U, Massachusetts General H, McGill U, National Eye Institute NIH. A global reference for human genetic variation. Nature 2015;526(7571):68–74.

