## Supplementary Document for "Pharmacogenomic study of heart failure and candesartan response from the CHARM programme"

**DATA SUPPLEMENT**

### Supplemental Results

##### *Functional annotation of the 8p21.3 region*

We identified 4 credible candidate variants within 500 kb of leading variant rs66886237 and with P-values within two orders of magnitude. The statistical prioritization performed using PAINTOR V3 failed to identify a marker with a high posterior probability to be causal (highest posterior probability=0.33). All variants share most of the functional annotations and therefore have similar deleteriousness scores. However, variant rs66886237 stands out as it ranked as the best candidate in RegulomeDB with a probability of being functional predicted at 0.61. While rs66886237 is in an intron of the *GFRA2* gene, we found no evidence of a functional impact on this gene and no strong associations were found between this variant and any traits or diseases.

We performed colocalization analysis on 126 different phenotypes (diseases and genes expression).[1-5] The colocalization analysis of GWAS of cardiovascular disease was concordant for the cardiomyopathy phenotype from in the FinnGen project (Freeze 4)[6] (H4=0.77, **Supplementary Figure 5**). With FinnGen, we found additional colocalizations driven by distinct signals including with coronary atherosclerosis (H3=0.96), major coronary heart disease event (H3=0.94), angina pectoris (H3=0.91) and ischemic heart diseases (H3=0.73); and with atrial fibrillation (H3=0.95) from the Roselli et al.[3] GWAS. We also found evidence of colocalization driven by the distinct signals for time-to-event phenotypes including myocardial infarction (H3=0.97) and other chronic ischemic heart disease (H3=0.96) in the UK Biobank.[7] We identified colocalizations driven by distinct signals in eQTL data for several genes including *DMTN*, *GFRA2* and *FAM160B2*. We queried the putative target genes in the ExPheWAS portal[8] to identify associations between phenotypes of the UK Biobank and the target genes using an approach that models the joint effect of variants. Those genes were found to be associated with gamma glutamyltransferase levels ($P=1.3\times10^{-50}$, $P=6.9\times10^{-13}$ and $P=8.3\times10^{-7}$ for *GFRA2*, *DOCK2* and *NPM2* respectively) and the mean corpuscular volume ($P=8.0\times10^{-10}$, $P=1.4\times10^{-6}$, $P=3.2\times10^{-62}$ and $P=6.7\times10^{-37}$ for *DMTN*, *GFRA2*, *DOCK2* and *NPM2* respectively) which are known to be associated with increased risk of cardiovascular disease.

##### *Functional annotation of the 3q31.13 region*

We identified 12 credible candidate variants within 500 kb of leading variant rs664669 at 3q13.13 and with P-values within two orders of magnitude. All variants share most of the functional annotations due to the high linkage disequilibrium, however, variant rs688405 stands out as it ranked as best candidate in DSNetwork and in RegulomeDB (ex quo with rs1152218). Variant rs688405 is located within a DNase I hypersensitive site in several tissues including cardiac fibroblast, is also located in a chromosomal interacting region that could be in contact with a distal gene *PVRL3* (also known as *NECTIN3*). PAINTOR identified rs201771335 as having a posterior probability to be causal of 1.0. Although the region of interest does not contain any known coding genes, the candidate variants are located within an intron of the long intergenic RNA, *RP11-457K10.1* which is mostly expressed in liver and kidney but also predicted as having coding potential in heart tissue (according to lncRNAKB)[9]. The nearest coding gene in the region is the developmental pluripotency associated 4 gene (*DPPA4),* which has genetic variants that were previously associated with congestive HF (beta= -0.049, P= $3.6\times10^{-6}$), diseases of the pericardium (beta= 0.004, P= $6.73\times10^{-7}$) in the UK Biobank.[7] It also has variants previously associated with endocarditis (OR= 2.9, P= $4.7\times10^{-6}$) in the FinnGen project (Freeze 4).[6] PhenoScanner identified associations between rs638409 and rs76356406 with cardiomegaly as cause of death from a study in the UK Biobank with p-values of $2.52\text{x}10^{-4}$ and $3.09\times10^{-4}$ respectively, located in the intron of RP11-457K10.1.

We found no colocalization driven by the same signal in both the CHARM analysis and the compared studies, with the highest H4 probabilities being 0.13 for cardiac arrest in FinnGen and 0.12 for *TRAT1* gene expression in whole blood. A single comparison, that with the *DPPA4* gene expression in whole blood, displays evidence of a colocalization driven by distinct signals (H3=0.94).

#### Supplemental Tables

***Supplementary Table 1.*** Summary information of the genetic data clean-up procedures performed prior to statistical analysis

| Clean-up step | N | Procedure | |
|---|---|---|---|
| | | SNP | IDs |
| Number of SNPs in genotyping file | 2,036,060 | | |
| Number of samples in genotyping file | 3,288 | | |
| Withdrawn individuals | 158 | | -158 |
| SNPs without physical position, Insertion/deletion variants and tri allelic variants removed | 58,318 | -58,318 | |
| Genotyping controls | 17 | | -17 |
| Failed markers | 279,977 | -279,977 | |
| Replicated SNPs excluded | 20,979 | -20,979 | |
| Samples with >10% missing genotypes | 7 | | -7 |
| SNPs with >2% missing genotypes | 1,352 | -1,352 | |
| Samples with >2% missing genotypes | 6 | | -6 |
| SNPs with plate-bias $P < 1 \cdot 10^{-7}$ | 291 | flagged | |
| SNPs used for IBS analysis | 74,896 | | |
| SNPs used for MDS analysis | 72,534 | | |
| Gender problem | 20 | | -20 |
| Discordant duplicated samples | 4 | | -4 |
| Related | 6 | | -6 |
| Ethnicity other than CEU by MDS cluster | 332 | | -332 |
| Haploid genotypes (after gender issues processed) | 509,373 | Set to missing | |
| SNPs with low completion rate on X chromosome | 406 | -406 | |
| SNPs with MAF=0 | 514,388 | -514,388 | |
| HWE test $4.4 \cdot 10^{-8} < P < 10^{-4}$ | 1,570 | flagged | |
| HWE test $P < 4.4 \cdot 10^{-8}$ (0.05/ 1,123,529) | 760 | -760 | |
| Final number of SNPs for analysis | 1,159,880 | | |
| Final number of samples for analysis | 2,738 | | |

***Supplementary Table 2.*** List of GWAS conducted

| GWAS # | Outcome | CHARM Study group | Variable Tested | Model | Study arms | n |
|---|---|---|---|---|---|---|
| **Primary HF Progression GWAS** | | | | | | |
| 1 | Composite CV endpoint | Overall | SNP | Cox | Both arms | 2727 |
| 2 | | Alt+Added | | | | 1698 |
| 3 | | Preserved | | | | 1029 |
| **Primary Candesartan Response GWAS** | | | | | | |
| 4 | Composite CV endpoint | Overall | SNP | Cox | Candesartan arm | 1371 |
| 5 | | Alt+Added | | | | 860 |
| 6 | | Preserved | | | | 511 |
| 7 | Hyperkalaemia | Overall | SNP | Logistic | Candesartan arm | 1371 |
| 8 | | Alt+Added | | | | 860 |
| 9 | | Preserved | | | | 511 |
| 10 | Renal dysfunction | Overall | | | | 1371 |
| 11 | | Alt+Added | | | | 860 |
| 12 | | Preserved | | | | 511 |
| 13 | Hypotension | Overall | | | | 1371 |
| 14 | | Alt+Added | | | | 860 |
| 15 | | Preserved | | | | 511 |
| 16 | Change SBP at week 6 post-treatment | Overall | SNP | GLM | Candesartan arm | 1285 |
| 17 | | Alt+Added | | | | 805 |
| 18 | | Preserved | | | | 480 |
| **Sensitivity/Exploratory Analysis GWAS** | | | | | | |
| 19 | Composite CV endpoint | Overall | SNP*Arm | Cox | Both arms | 2727 |
| 20 | | Alt+Added | | | | 1698 |
| 21 | | Preserved | | | | 1029 |
| 22 | Hyperkalaemia | Overall | SNP*Arm | Logistic | Both arms | 2727 |
| 25 | | Alt+Added | | | | 1698 |
| 28 | | Preserved | | | | 1029 |
| 23 | Renal dysfunction | Overall | | | | 2727 |
| 26 | | Alt+Added | | | | 1698 |
| 29 | | Preserved | | | | 1029 |
| 24 | Hypotension | Overall | | | | 2727 |
| 27 | | Alt+Added | | | | 1698 |
| 30 | | Preserved | | | | 1029 |
| 31 | Change SBP at week 6 post-treatment (adjusted for baseline) | Overall | SNP | GLM | Candesartan arm | 1285 |
| 32 | | Alt+Added | | | | 805 |
| 33 | | Preserved | | | | 480 |
| 34 | | Overall | SNP*Arm | GLM | Both arms | 2561 |
| 35 | | Alt+Added | | | | 1579 |
| 36 | | Preserved | | | | 982 |
| 37 | Change SBP at week 6 post-treatment | Overall | | | | 2561 |
| 38 | | Alt+Added | | | | 1579 |
| 39 | | Preserved | | | | 982 |

***Supplementary Table 3.*** Descriptive statistics of the CHARM-Preserved study according to genetic variant rs66886237 genotypes at 8p21.3

| | **rs66886237** | | |
|---|---|---|---|
| | **GG** | **GA** | **AA** |
| **Number of patients, n** | **751** | **251** | **22** |
| Age, mean ± SD | 67.5±10.6 | 68.7±11.2 | 70.0±9.4 |
| Men, n (%) | 419 (55.8%) | 130 (51.8%) | 12 (54.6%) |
| European origin, n (%) | 720 (95.9%) | 237 (94.4%) | 22 (100%) |
| ***Heart disease risk factors*** | | | |
| NYHA class II, n (%) | 420 (55.9%) | 126 (50.2%) | 13 (59.1%) |
| NYHA class III, n (%) | 314 (41.8%) | 121 (48.2%) | 9 (40.9%) |
| NYHA class IV, n (%) | 17 (2.3%) | 4 (1.6%) | 0 (0%) |
| LVEF, mean ± SD | 0.5±0.1 | 0.5±0.1 | 0.5±0.1 |
| BMI (kg/m2), mean ± SD | 29.9±6.3 | 29.9±6.2 | 28.2±4.9 |
| Heart rate (beats/min), mean ± SD | 70.4±12.2 | 72.1±13.3 | 74.0±10.8 |
| Systolic blood pressure (mm HG), mean ± SD | 134.6±18.9 | 133.3±19.0 | 135.0±20.0 |
| Diastolic blood pressure (mm Hg), mean ± SD | 76.3±10.7 | 74.5±11.2 | 75.7±9.6 |
| Current smoker, n (%) | 97 (12.9%) | 34 (13.6%) | 2 (9.1%) |
| ***Heart failure cause*** | | | |
| Ischemic HF, n (%) | 388 (51.7%) | 137 (54.6%) | 14 (63.6%) |
| Idiopathic HF, n (%) | 84 (11.2%) | 30 (12.0%) | 3 (13.6%) |
| Hypertensive HF, n (%) * | 193 (25.7%) | 48 (19.1%) | 1 (4.6%) |
| ***Medical history prior to baseline*** | | | |
| Myocardial infarction, n (%) | 327 (43.5%) | 113 (45.0%) | 7 (31.8%) |
| Angina, n (%) | 417 (55.5%) | 139 (55.4%) | 13 (59.1%) |
| Stroke, n (%) | 78 (10.4%) | 30 (12.0%) | 1 (4.6%) |
| Diabetes, n (%) | 221 (29.4%) | 85 (33.9%) | 4 (18.2%) |
| Hypertension, n (%) | 502 (66.8%) | 178 (70.9%) | 12 (54.6%) |
| Atrial fibrillation at baseline, n (%) | 234 (31.2%) | 91 (36.3%) | 7 (31.8%) |
| PCI, n (%) * | 114 (15.2%) | 47 (18.7%) | 10 (45.5%) |
| CABG, n (%) | 174 (23.2%) | 61 (24.3%) | 8 (36.4%) |
| ***Medication at baseline*** | | | |
| ACE inhibitors, n (%) | 150 (20.0%) | 52 (20.7%) | 3 (13.6%) |
| Diuretics, n (%) | 601 (80.0%) | 206 (82.1%) | 18 (81.8%) |
| B-blocker, n (%) | 415 (55.3%) | 134 (53.4%) | 10 (45.5%) |
| Spironolactone, n (%) | 121 (16.1%) | 33 (13.2%) | 3 (13.6%) |
| Digoxin/digitalis glycoside, n (%) | 237 (31.6%) | 84 (33.5%) | 5 (22.7%) |
| Aspirin, n (%) | 428 (57.0%) | 147 (58.6%) | 11 (50.0%) |
| Other antiplatelet agents, n (%) | 26 (3.5%) | 12 (4.8%) | 3 (13.6%) |
| Lipid-lowering drug, n (%) | 333 (44.3%) | 109 (43.4%) | 12 (54.6%) |
| ***Study outcomes*** | | | |
| CV death or HF hospit., n (%) * | 160 (21.3%) | 99 (39.4%) | 11 (50.0%) |
| CV death, n (%) * | 62 (8.3%) | 47 (18.7%) | 5 (22.7%) |
| Any cause death, n (%) * | 103 (13.7%) | 59 (23.5%) | 6 (27.3%) |
| Hypotension, n (%) | 85 (11.3%) | 42 (16.7%) | 3 (13.6%) |

| | | | |
|---|---|---|---|
| Renal dysfunction, n (%) | 85 (11.3%) | 36 (14.3%) | 3 (13.6%) |
| Hyperkalaemia, n (%) | 24 (3.2%) | 11 (4.4%) | 0 (0%) |

ACE: angiotensin converting enzyme; BMI: body mass indez; CABG: coronary artery bypass graft; CV: cardiovascular; HF: heart failure; LVEF: left ventricular ejection fraction; N: number of patients; NYHA: New York Heart Association; PCI: percutaneous coronary intervention; SD: standard deviation.

* P<0.01 according to non parametric test (Kruscal-Wallis, Chi squared or Fisher exact test)

***Supplementary Table 4.*** Association results for the composite of cardiovascular death or heart failure hospitalisation in the CHARM studies for the leading genetic variant rs664669 at 3q13.13

| **Genetic variant :effect allele** | **CHARM studies** | **Study arm** | **N** | **N Events (%)** | **HR (95% CI)** | **P value** | **Interaction P value*** |
|---|---|---|---|---|---|---|---|
| rs664669:C | Overall | Candesartan | 1369 | 435 (31.8%) | 1.48 (1.29-1.69) | $2.63\times10^{-8}$ | 0.003 |
| | | Placebo | 1353 | 481 (35.6%) | 1.11 (0.97-1.26) | 0.125 | |
| | Alternative | Candesartan | 381 | 117 (30.7%) | 1.69 (1.29-2.21) | $1.21\times10^{-4}$ | 0.022 |
| | | Placebo | 373 | 154 (41.3%) | 1.06 (0.84-1.35) | 0.626 | |
| | Added | Candesartan | 478 | 185 (38.7%) | 1.55 (1.25-1.93) | $6.33\times10^{-5}$ | 0.077 |
| | | Placebo | 462 | 189 (40.9%) | 1.15 (0.93-1.42) | 0.195 | |
| | Alternative + Added | Candesartan | 859 | 302 (35.2%) | 1.59 (1.35-1.88) | $4.70\times10^{-8}$ | 0.004 |
| | | Placebo | 835 | 343 (41.1%) | 1.11 (0.95-1.30) | 0.184 | |
| | Preserved | Candesartan | 510 | 133 (26.1%) | 1.33 (1.03-1.70) | 0.026 | 0.266 |
| | | Placebo | 518 | 138 (26.6%) | 1.12 (0.88-1.42) | 0.368 | |

CI: confidence interval; HR: hazard ratio; N: number of patients

*Interaction P value for the genetic variant by treatment arm interaction term in a Cox regression model

***Supplementary Table 5.*** Effect of candesartan on the composite cardiovascular endpoint of CV death or hospitalisation for heart failure in the CHARM studies, according to genetic subgroups for the leading genetic variant rs664669 at 3q13.13**.**

| | | | | **Total** | | **Candesartan** | | **Placebo** | | | |
|---|---|---|---|---|---|---|---|---|---|---|---|
| | **Study** | | **Group %** | **N total** | **N events (%)** | **N total** | **N events (%)** | **N total** | **N events (%)** | **HR (95% CI)** | **P value** |
| rs664669 | OVERALL | C/C | 19.0% | 518 | 215 (41.5%) | 247 | 104 (42.1%) | 271 | 111 (41.0%) | 1.00 (0.76-1.31) | 0.983 |
| | | C/T | 49.6% | 1350 | 457 (33.9%) | 699 | 235 (33.6%) | 651 | 222 (34.1%) | 0.97 (0.81-1.17) | 0.769 |
| | | T/T | 31.4% | 854 | 244 (28.6%) | 423 | 96 (22.7%) | 431 | 148 (34.3%) | 0.59 (0.45-0.76) | 6.13E-05 |
| | ALTERNATIVE | C/C | 16.2% | 122 | 57 (46.7%) | 59 | 25 (42.4%) | 63 | 32 (50.8%) | 0.78 (0.45-1.36) | 0.380 |
| | | C/T | 50.1% | 378 | 135 (35.7%) | 192 | 66 (34.4%) | 186 | 69 (37.1%) | 0.88 (0.63-1.25) | 0.487 |
| | | T/T | 33.7% | 254 | 79 (31.1%) | 130 | 26 (20.0%) | 124 | 53 (42.7%) | 0.37 (0.23-0.60) | 7.01E-05 |
| | ADDED | C/C | 19.9% | 187 | 90 (48.1%) | 90 | 44 (48.9%) | 97 | 46 (47.4%) | 1.21 (0.77-1.91) | 0.407 |
| | | C/T | 50.5% | 475 | 190 (40.0%) | 249 | 102 (41.0%) | 226 | 88 (38.9%) | 1.09 (0.81-1.46) | 0.581 |
| | | T/T | 29.6% | 278 | 94 (33.8%) | 139 | 39 (28.1%) | 139 | 55 (39.6%) | 0.66 (0.43-1.01) | 0.055 |
| | ALTERNATIVE + ADDED | C/C | 18.2% | 309 | 147 (47.6%) | 149 | 69 (46.3%) | 160 | 78 (48.8%) | 0.96 (0.68-1.35) | 0.829 |
| | | C/T | 50.4% | 853 | 325 (38.1%) | 441 | 168 (38.1%) | 412 | 157 (38.1%) | 1.00 (0.81-1.25) | 0.973 |
| | | T/T | 31.4% | 532 | 173 (32.5%) | 269 | 65 (24.2%) | 263 | 108 (41.1%) | 0.52 (0.38-0.71) | 3.90E-05 |
| | PRESERVED | C/C | 20.3% | 209 | 68 (32.5%) | 98 | 35 (35.7%) | 111 | 33 (29.7%) | 1.18 (0.71-1.95) | 0.521 |
| | | C/T | 48.3% | 497 | 132 (26.6%) | 258 | 67 (26.0%) | 239 | 65 (27.2%) | 0.91 (0.65-1.30) | 0.617 |
| | | T/T | 31.3% | 322 | 71 (22.0%) | 154 | 31 (20.1%) | 168 | 40 (23.8%) | 0.82 (0.50-1.35) | 0.442 |

CI: confidence interval; CV: cardiovascular; HR: hazard ratio; N: number of patients

***Supplementary Table 6.*** Gene level collapsing models

| **Qualifying variant model** | **External GnomAD MAF thresholds** | **Internal MAF thresholds** | **Variant consequence impact** | **Missense predicted damaging** | **Missense tolerance ratio (MTR) score** |
|---|---|---|---|---|---|
| Synonymous | ≤ 0.005% (global) | ≤ 0.05% | Synonymous<br>(Negative Control) | - | - |
| Protein-truncating variants (PTV) | ≤ 0.1% (global and popmax) | ≤ 0.1% | PTV | - | - |
| Ultra-Rare Damaging | not in GnomAD | ≤ 0.025% | PTV, missense, inframe indels | REVEL ≥ 0.25 | - |
| Ultra-Rare Damaging + MTR | not in GnomAD | ≤ 0.025% | PTV, missense, inframe indels | REVEL ≥ 0.25 | MTR ≤ 25%ile OR<br>Intragenic MTR ≤ 50%ile |
| Rare damaging | ≤ 0.005% (global) | ≤ 0.05% | PTV, missense, inframe indels | REVEL ≥ 0.25 | - |
| Rare damaging + MTR | ≤ 0.005% (global) | ≤ 0.05% | PTV, missense, inframe indels | REVEL ≥ 0.25 | MTR ≤ 25%ile OR<br>Intragenic MTR ≤ 50%ile |
| Flexible damaging | ≤ 0.05% (global)<br>≤ 0.1% (popmax) | ≤ 0.1% | PTV, missense, inframe indels | REVEL ≥ 0.25 | - |
| Flexible non-synonymous | ≤ 0.05% (global)<br>≤ 0.1% (popmax) | ≤ 0.1% | PTV, missense, inframe indels | - | - |
| Flexible non-synonymous + MTR | ≤ 0.05% (global)<br>≤ 0.1% (popmax) | ≤ 0.1% | PTV, missense, inframe indels | - | MTR ≤ 25%ile OR<br>Intragenic MTR ≤ 50%ile |
| Protein-truncating variant or rare damaging missense | PTV ≤ 0.1% (global and popmax)<br>missense ≤ 0.005% (global)<br>missense ≤ 0.05% (popmax) | PTV ≤ 0.1%<br>missense ≤ 0.05% | PTV, missense, inframe indels | REVEL ≥ 0.25 | - |
| Recessive Non-Synonymous | ≤ 0.5% (global and popmax) | ≤ 0.5% | PTV, missense, inframe indels, AND variant homozygous OR two heterozygous variants in same gene | - | - |

MAF: minor allele frequency: MTR: missense tolerance ratio; PTV: protein truncating variant. The following parameters adopted universally to all models: Minimum coverage 10X; Has annotations in CCDS transcripts (CCDS release 22; ~34Mb); Percent alternate reads in homozygous variants >= 0.8; Percent alternate reads in heterozygous variants >= 0.3 and <= 0.8; Binomial test of alternate reads proportion p > 0.000001; Genotype quality score (GQ) >= 30; Fisher's strand bias score (FS) <= 200 (indels) <= 60 (SNVs); Mapping quality score (MQ) >= 40; Quality score (QUAL) >= 30; Read position rank sum score (RPRS) >= -2; Mapping quality rank sum score (MQRS) >= -8; DRAGEN variant status = PASS; Binomial test of difference in coverage between cases and controls p 0.000001; Variant site achieved 10-fold coverage in >=25% of GnomAD samples, and if variant was observed in GnomAD the variant calls in GnomAD achieved exome z score >=2.0, exome MQ >= 30 and exome allele count raw percent >= 50; Not in a list of 951 observed sequencing artefacts or problematic variants.

***Supplementary Table 7.*** Genome-wide gene-level collapsing analysis top results

**a.** Reporting association results with the endpoint of cardiovascular death or heart failure hospitalisation tested in 1010 patients in both arms of the CHARM-Preserved study, adjusted for age, treatment arm, and sex.

| | | **Gene** | | | **Age** | | **Sex** | | **Treatment** | |
|---|---|---|---|---|---|---|---|---|---|---|
| **Model*** | **Gene** | **N carriers** | **Beta** | **P-value** | **Beta** | **P-value** | **Beta** | **P-value** | **Beta** | **P-value** |
| Flexible damaging | *MCHR1* | 5 | 3.990 | 1.13E-04 | 0.043 | 2.98E-09 | 0.033 | 8.24E-01 | -0.073 | 6.18E-01 |
| | *HELQ* | 7 | 2.842 | 3.32E-04 | 0.042 | 4.66E-09 | 0.038 | 7.98E-01 | -0.060 | 6.81E-01 |
| | *TAS1R1* | 12 | 2.143 | 3.34E-04 | 0.041 | 1.08E-08 | 0.068 | 6.46E-01 | -0.077 | 6.00E-01 |
| | *TRIM45* | 9 | 2.422 | 3.46E-04 | 0.043 | 3.92E-09 | 0.079 | 5.94E-01 | -0.087 | 5.54E-01 |
| | *SLC5A2* | 11 | 2.094 | 6.44E-04 | 0.042 | 8.49E-09 | 0.020 | 8.95E-01 | -0.050 | 7.35E-01 |
| Flexible non- synonymous | *MCHR1* | 8 | 3.043 | 6.01E-05 | 0.043 | 3.22E-09 | 0.033 | 8.26E-01 | -0.080 | 5.85E-01 |
| | *SERTAD2* | 8 | 2.953 | 1.01E-04 | 0.042 | 4.61E-09 | 0.061 | 6.83E-01 | -0.081 | 5.83E-01 |
| | *SMPD2* | 5 | 3.516 | 5.30E-04 | 0.041 | 1.05E-08 | 0.062 | 6.75E-01 | -0.075 | 6.09E-01 |
| | *SLC5A2* | 11 | 2.094 | 6.44E-04 | 0.042 | 8.49E-09 | 0.020 | 8.95E-01 | -0.050 | 7.35E-01 |
| | *NOTCH4* | 33 | 1.230 | 7.65E-04 | 0.042 | 7.33E-09 | 0.052 | 7.26E-01 | -0.070 | 6.33E-01 |
| | *PDE10A* | 5 | 3.530 | 7.77E-04 | 0.041 | 1.80E-08 | 0.030 | 8.40E-01 | -0.090 | 5.37E-01 |
| | *KLC1* | 7 | 2.630 | 8.30E-04 | 0.041 | 9.88E-09 | 0.047 | 7.51E-01 | -0.047 | 7.47E-01 |
| Flexible non- synonymous + MTR | *MTCL1* | 19 | 2.163 | 9.67E-06 | 0.039 | 8.01E-08 | 0.066 | 6.58E-01 | -0.064 | 6.66E-01 |
| | *TNFRSF25* | 5 | 3.708 | 3.60E-04 | 0.042 | 8.22E-09 | 0.030 | 8.42E-01 | -0.057 | 6.95E-01 |
| | *NOTCH4* | 16 | 1.781 | 5.75E-04 | 0.042 | 4.98E-09 | 0.014 | 9.25E-01 | -0.046 | 7.52E-01 |
| | *VPS13C* | 23 | -2.883 | 7.95E-04 | 0.041 | 1.03E-08 | 0.064 | 6.68E-01 | -0.067 | 6.48E-01 |
| | *ACAP3* | 9 | 2.265 | 9.05E-04 | 0.041 | 8.44E-09 | 0.083 | 5.77E-01 | -0.051 | 7.29E-01 |
| Protein-truncating variant or rare damaging missense | *NOTCH4* | 12 | 1.976 | 7.90E-04 | 0.042 | 3.94E-09 | 0.050 | 7.37E-01 | -0.056 | 7.03E-01 |
| Rare damaging + MTR | *MTCL1* | 9 | 2.584 | 4.09E-04 | 0.039 | 4.68E-08 | 0.067 | 6.52E-01 | -0.049 | 7.39E-01 |
| | *C16orf96* | 6 | 3.408 | 4.99E-04 | 0.040 | 3.12E-08 | 0.081 | 5.89E-01 | -0.068 | 6.43E-01 |

**b.** Reporting association results with the endpoint of cardiovascular death or heart failure hospitalisation tested in 1346 patients in the candesartan treatment arms of CHARM-Overall, adjusted for age and sex.

| | | Gene | | | Age | | Sex | |
|---|---|---|---|---|---|---|---|---|
| **Model*** | **Gene** | **N carriers** | **Beta** | **P-value** | **Beta** | **P-value** | **Beta** | **P-value** |
| Flexible damaging | *RASSF7* | 6 | 3.295 | 8.56E-04 | 0.026 | 4.01E-06 | -0.424 | 1.05E-03 |
| | *TRPV2* | 8 | 2.468 | 9.58E-04 | 0.027 | 1.90E-06 | -0.420 | 1.19E-03 |
| Flexible non- synonymous | *STRN3* | 13 | 2.313 | 1.11E-04 | 0.026 | 5.15E-06 | -0.439 | 7.22E-04 |
| | *KDM2A* | 7 | 3.543 | 1.57E-04 | 0.027 | 1.79E-06 | -0.412 | 1.48E-03 |
| | *PRMT9* | 11 | 2.572 | 1.88E-04 | 0.026 | 5.94E-06 | -0.398 | 2.11E-03 |
| | *SHKBP1* | 12 | 2.067 | 5.62E-04 | 0.028 | 7.28E-07 | -0.422 | 1.11E-03 |
| | *CRISP1* | 5 | 3.585 | 6.39E-04 | 0.028 | 8.85E-07 | -0.422 | 1.10E-03 |
| | *PPP1R3F* | 5 | 3.523 | 6.81E-04 | 0.028 | 1.21E-06 | -0.434 | 8.14E-04 |
| | *MGAT1* | 6 | 3.327 | 7.87E-04 | 0.026 | 2.91E-06 | -0.414 | 1.36E-03 |
| | *MYOD1* | 10 | 2.174 | 8.43E-04 | 0.027 | 2.32E-06 | -0.448 | 5.50E-04 |
| Protein-truncating variant or rare damaging missense | *TRPV2* | 8 | 2.468 | 9.58E-04 | 0.027 | 1.90E-06 | -0.420 | 1.19E-03 |
| Protein-truncating variants | *ERN2* | 7 | 3.443 | 2.66E-04 | 0.026 | 3.74E-06 | -0.421 | 1.15E-03 |
| Rare damaging | *TRPV2* | 8 | 2.468 | 9.58E-04 | 0.027 | 1.90E-06 | -0.420 | 1.19E-03 |
| Recessive Non-Synonymous | *NPHP4* | 5 | 3.496 | 5.98E-04 | 0.028 | 1.07E-06 | -0.432 | 8.44E-04 |
| | *MXRA5* | 19 | 1.604 | 6.42E-04 | 0.028 | 1.06E-06 | -0.395 | 2.28E-03 |
| Ultra-Rare Damaging + MTR | *MCPH1* | 20 | -3.000 | 4.40E-04 | 0.026 | 3.20E-06 | -0.437 | 7.27E-04 |

* Models are described in Supplementary Table 6.

***Supplementary Table 8.*** Gene-level collapsing analysis results in candidate regions from GWAS

**a.** Reporting P-values (number of carriers) for genes on chr8:20,599,236-22,607,852 for the endpoint of CV death or hospitalisation for HF tested in 1,010 patients in both arms of the CHARM-Preserved study, adjusted for age, treatment arm, and sex.

| **Position** | **Gene** | **Flexible non-synonymous** | **Flexible non-synonymous + MTR** | **Flexible damaging** | **PTV or rare damaging missense** | **Rare damaging** | **Rare damaging + MTR** | **PTV** | **Recessive non-synonymous** | **Ultra-Rare Damaging** | **Ultra-Rare Damaging + MTR** | **Synonymous (control)** |
|---|---|---|---|---|---|---|---|---|---|---|---|---|
| 21550790 | *GFRA2* | 0.43 (8) | 0.56 (5) | 0.74 (4) | 0.99 (3) | 0.99 (3) | 0.99 (3) | 0.81 (2) | - | - | - | - |
| 21766822 | *DOK2* | 0.11 (10) | 0.17 (6) | 0.08 (7) | 0.67 (3) | 0.67 (3) | - | - | - | - | - | 0.59 (3) |
| 21777282 | *XPO7* | - | - | - | - | - | - | - | - | - | - | 0.91 (4) |
| 21882760 | *NPM2* | 0.39 (5) | 0.22 (2) | 0.22 (2) | - | - | - | - | - | - | - | 0.71 (2) |
| 21924387 | *DMTN* | 0.58 (7) | 0.73 (2) | - | - | - | - | - | - | - | - | - |
| 21946765 | *FAM160B2* | 0.77 (9) | 0.37 (7) | 0.8 (2) | 0.8 (2) | 0.8 (2) | 0.8 (2) | - | - | - | - | 0.25 (2) |
| 21964811 | *NUDT18* | 0.71 (2) | 0.71 (2) | | | | | - | - | - | - | 0.56 (2) |
| 21973213 | *HR* | 0.92 (15) | 0.61 (9) | 0.74 (7) | 0.89 (6) | 0.89 (6) | 0.87 (4) | - | - | - | - | 0.59 (4) |
| 21995863 | *REEP4* | 0.51 (2) | | | | | | - | - | - | - | |
| 22005673 | *LGI3* | 0.08 (7) | 0.02 (3) | 0.08 (7) | 0.24 (4) | 0.24 (4) | | - | - | - | - | 0.64 (2) |
| 22019342 | *SFTPC* | 0.25 (2) | | | | | | - | - | - | - | |
| 22022919 | *BMP1* | 0.9 (14) | 0.64 (7) | 0.65 (7) | 0.64 (3) | 0.64 (3) | 0.64 (3) | - | - | 0.97 (2) | 0.97 (2) | 0.31 (10) |
| 22078866 | *PHYHIP* | - | - | - | - | - | - | - | - | - | - | 0.27 (2) |
| 22102963 | *POLR3D* | 0.25 (6) | - | - | - | - | - | - | - | - | - | - |
| 22136900 | *PIWIL2* | 0.79 (10) | 0.63 (7) | 0.76 (5) | 0.97 (4) | 0.97 (4) | 0.79 (3) | - | - | 0.6 (2) | 0.6 (2) | 0.42 (7) |
| 22262224 | *SLC39A14* | 0.3 (6) | 0.81 (3) | 0.08 (2) | | | - | - | - | - | - | - |
| 22298923 | *PPP3CC* | 0.7 (4) | 0.54 (3) | 0.82 (2) | 0.82 (2) | 0.82 (2) | - | - | - | - | - | - |
| 22412020 | *SORBS3* | 0.36 (12) | 0.69 (7) | - | - | - | - | 0.64 (3) | 0.87 (2) | - | - | 0.39 (2) |
| 22436293 | *PDLIM2* | 0.55 (7) | 0.51 (3) | 0.89 (2) | - | - | - | - | - | - | - | - |
| 22457234 | *C8orf58* | 0.32 (7) | 0.77 (3) | - | - | - | - | - | - | - | - | 0.83 (3) |
| 22463287 | *CCAR2* | 0.66 (17) | 0.34 (9) | 0.58 (3) | - | - | - | - | - | - | - | 0.61 (3) |
| 22478935 | *BIN3* | 0.38 (10) | 0.71 (9) | 0.79 (6) | - | - | - | - | - | - | - | - |

| | | | | | | | | | | | | |
|---|---|---|---|---|---|---|---|---|---|---|---|---|
| 22547986 | *EGR3* | 0.47 (3) | - | - | - | - | - | - | - | - | - | - |
| 22570883 | *PEBP4* | 0.86 (5) | 0.53 (3) | 0.74 (2) | 0.74 (2) | - | - | 0.59 (2) | - | - | - | - |

MTR: Missense tolerance ratio; PTV: Protein truncating variant; Positions in build hg37.

**b.** Reporting P-values (number of carriers) for genes on chr3:107,476,144-111,503,214 for the endpoint of cardiovascular death or heart failure hospitalisation tested in 1,346 patients in the candesartan treatment arm of CHARM-Overall, adjusted for age and sex.

| **Position** | **Gene** | **Flexible non-synonymous** | **Flexible non-synonymous + MTR** | **Flexible damaging** | **PTV or rare damaging missense** | **Rare damaging** | **Rare damaging + MTR** | **PTV** | **Ultra-Rare Damaging** | **Ultra-Rare Damaging + MTR** | **Synonymous (control)** |
|---|---|---|---|---|---|---|---|---|---|---|---|
| 107766131 | *CD47* | 0.33 (4) | 0.77 (2) | - | - | - | - | - | - | - | 0.11 (3) |
| 107881324 | *IFT57* | 0.62 (3) | 0.78 (3) | 0.10 (3) | 0.40 (2) | 0.40 (2) | 0.40 (2) | - | - | - | 0.66 (1) |
| 108047151 | *HHLA2* | 0.76 (3) | 0.91 (2) | - | - | - | - | - | - | - | 0.25 (4) |
| 108100392 | *MYH15* | 0.41 (37) | 0.34 (20) | 0.93 (16) | 0.87 (11) | 0.87 (11) | 0.68 (6) | - | 0.19 (6) | 0.12 (5) | 0.33 (9) |
| 108269996 | *CIP2A* | 0.48 (14) | 0.70 (7) | 0.97 (5) | 0.79 (4) | 0.77 (4) | 0.77 (4) | 0.77 (2) | 0.58 (1) | 0.58 (1) | - |
| 108324254 | *DZIP3* | 0.92 (14) | 0.23 (9) | 0.31 (2) | 0.31 (2) | - | - | - | - | - | 0.006 (4) |
| 108474625 | *RETNLB* | 0.45 (3) | - | 0.68 (2) | - | - | - | - | - | - | - |
| 108541775 | *TRAT1* | 0.44 (3) | - | - | - | - | - | - | - | - | - |
| 108626869 | *GUCA1C* | 0.92 (3) | 0.86 (2) | - | - | - | - | - | - | - | - |
| 108677812 | *MORC1* | 0.42 (6) | 0.49 (2) | - | - | - | - | - | - | - | 0.79 (2) |
| 109019240 | *DPPA2* | 0.65 (9) | 0.51 (6) | - | - | - | - | - | - | - | 0.69 (2) |
| 109046835 | *DPPA4* | 0.55 (5) | - | - | - | - | - | - | - | - | - |
| 110790865 | *NECTIN3* | 0.61 (15) | 0.43 (12) | 0.81 (4) | 0.81 (4) | 0.81 (4) | 0.22 (3) | - | - | - | 0.30 (3) |
| 111261096 | *CD96* | 0.50 (10) | 0.25 (4) | 0.68 (2) | 0.68 (2) | 0.68 (2) | - | - | - | - | 0.52 (2) |
| 111312392 | *ZBED2* | 0.98 (2) | - | - | - | - | - | - | - | - | - |
| 111394093 | *PLCXD2* | 0.38 (8) | 0.89 (2) | 0.83 (2) | 0.83 (2) | 0.83 (2) | - | - | - | - | - |

MTR: Missense tolerance ratio; PTV: Protein truncating variant; Positions in build hg37.

#### Supplemental Figures

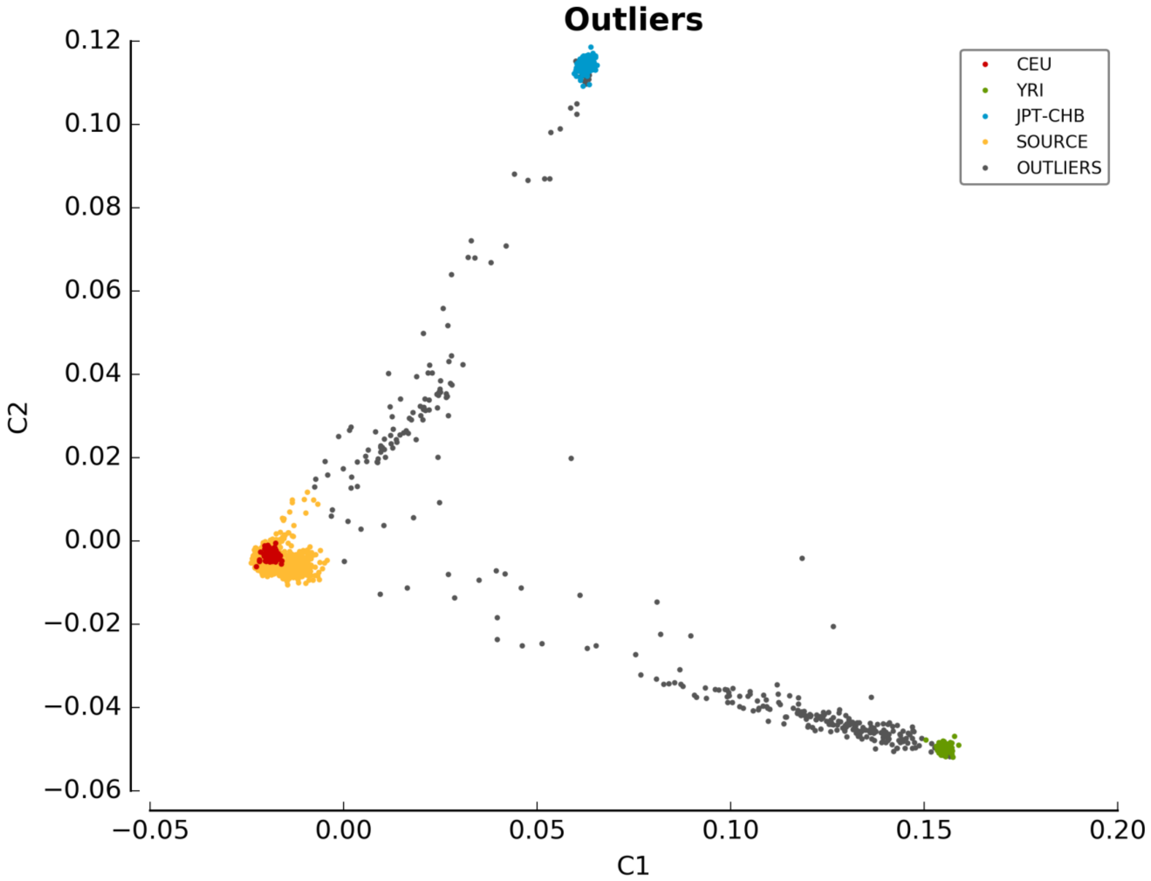


***Supplementary Figure 1.*** MDS plots for genetic ancestry

MDS plots showing the first two principal components of the source dataset with the reference panels. The outliers of the CEU population are shown in grey, while samples of the source dataset that resemble the CEU population is shown in orange. A multiplier of 1.9 was used to find the 332 outliers.

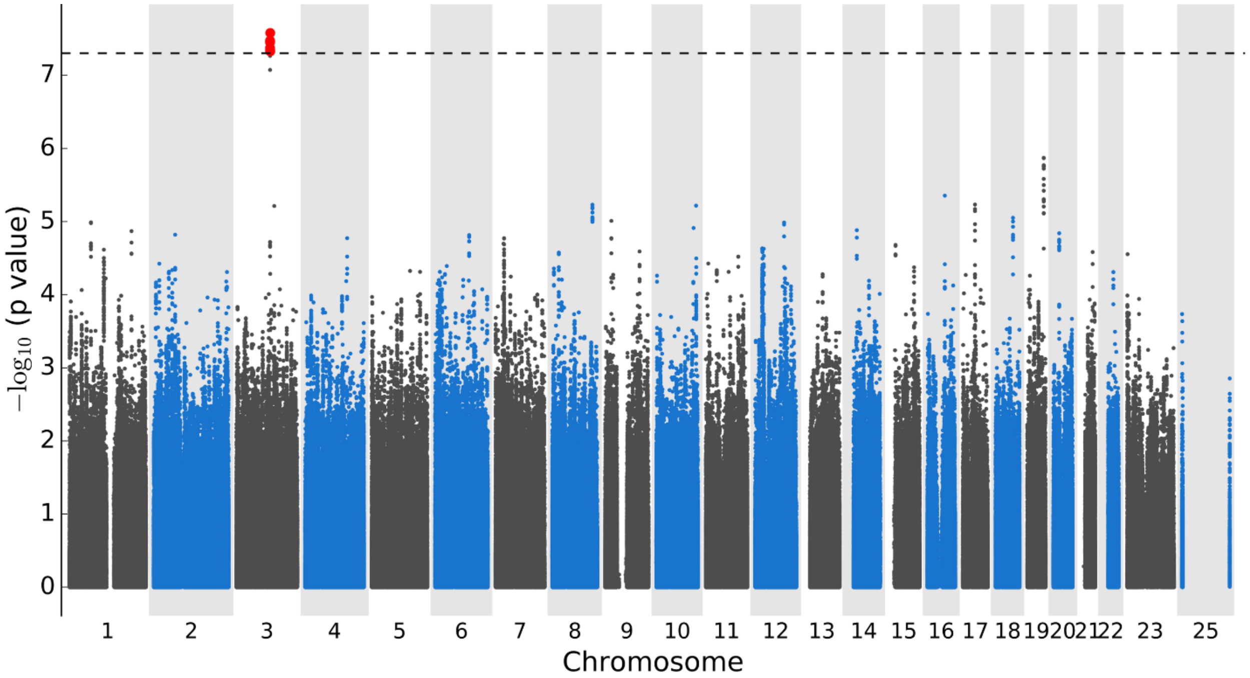


***Supplementary Figure 2.*** Manhattan plot of the GWAS with 1371 patients from the candesartan arms of the CHARM-Overall program tested for association with time to cardiovascular death or heart failure hospitalisation using Cox proportional hazards regression with adjustment for principal components for genetic ancestry, age, and sex. There were 5,015,431 genotyped and imputed genetic variants of MAF ≥ 5%. The dashed line marks $P=5\times10^{-8}$.

***Supplementary Figure 3. Regional plots***

The first y-axis shows the negative $\log_{10}$ of P values for genotyped (circles and darker colors) and imputed (lozenges and paler colors) genetic variants, the second y-axis shows the recombination rate from HapMap reference samples (black line). Genes are displayed below the graph, base pair positions are given according to hg19, the degree of linkage disequilibrium ($r^2$) of each genetic variant with the top genetic variant estimated from the study population is displayed as blue for [0, 0.2], purple for [0.2, 0.4], green for [0.4, 0.6], orange for [0.6, 0.8], and red for [0.8, 1.0]. Gene information from Ensembl (build37). The dashed line marks $P=5\times10^{-8}$.

**a.** Chromosome 8 region (21,107,231-22,107,231) for association with time to cardiovascular death or heart failure hospitalisation in 1029 patients from the CHARM-Preserved study.

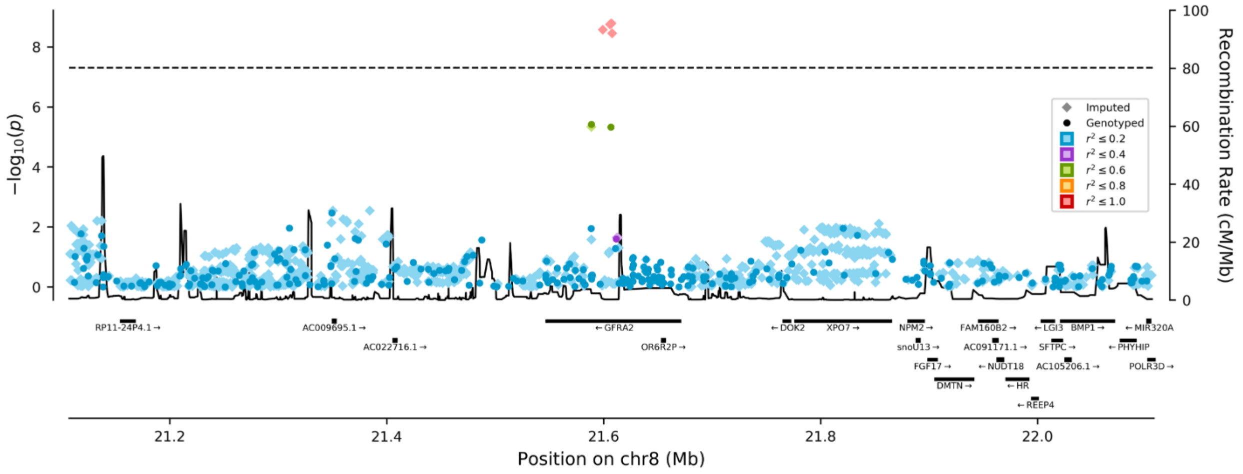


**b.** Chromosome 3 region (109,001,313-110,001,313) for association with time to cardiovascular death or heart failure hospitalisation in 1371 patients from the candesartan arms of the CHARM-Overall program.

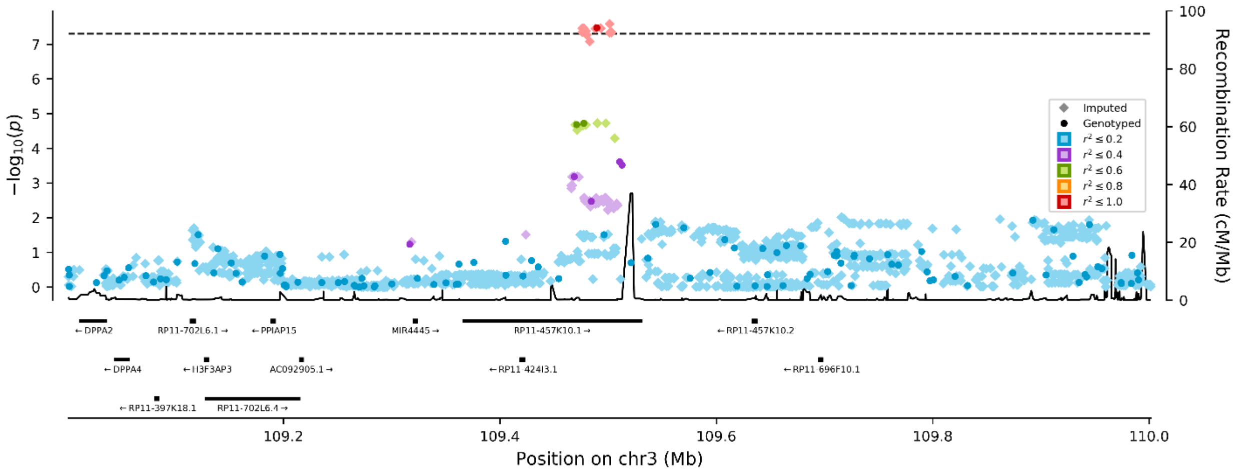

*Supplementary Figure 4. QQ plots*

**a.** QQ plot of the P values of the GWAS with 1029 heart failure patients with preserved ejection fraction from the CHARM-Preserved study for 5,023,375 genotyped and imputed genome-wide common genetic variants (MAF ≥ 5%) tested for association with time to cardiovascular death or heart failure hospitalisation using Cox proportional hazards regression (inflation factor λ = 1.01).

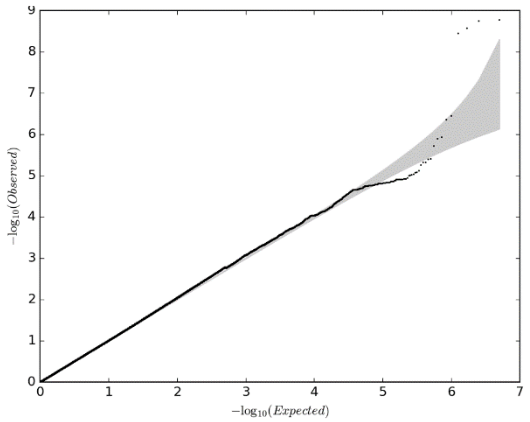


**b.** QQ plot of the P values of the GWAS with 1371 heart failure patients from the candesartan arms of the CHARM-Overall program for 5,015,431 genotyped and imputed genome-wide common genetic variants (MAF ≥ 5%) tested for association with time to cardiovascular death or heart failure hospitalisation using Cox proportional hazards regression (inflation factor λ = 1.01).

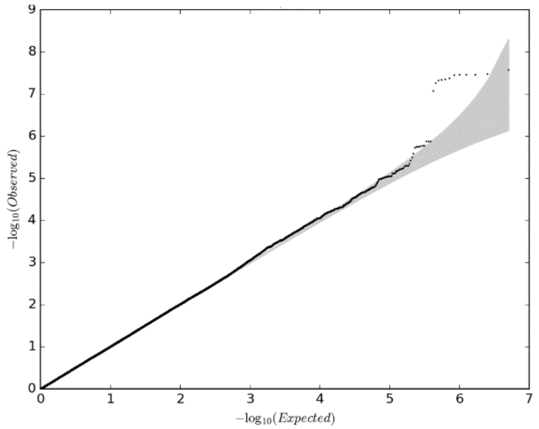

***Supplementary Figure 5.*** P values QQ plot for genome-wide gene-level collapsing analysis.

**a.** Showing results of the synonymous model (negative control) cardiovascular death or heart failure hospitalisation using Firth's logistic regression with 1010 patients in CHARM-Preserved, controlling for sex, age, treatment arm (inflation factor λ = 0.7927).

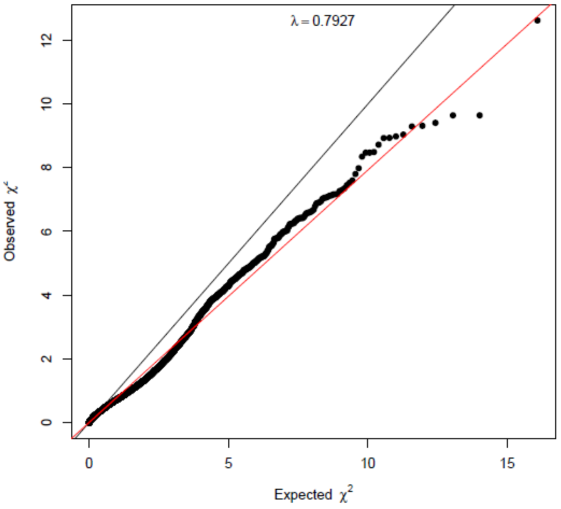


**b.** Showing results of the synonymous model (negative control) for cardiovascular death or heart failure hospitalisation with 1346 patients in the candesartan treatment arm of CHARM-Overall, controlling for age and sex (inflation factor λ = 0.7928).

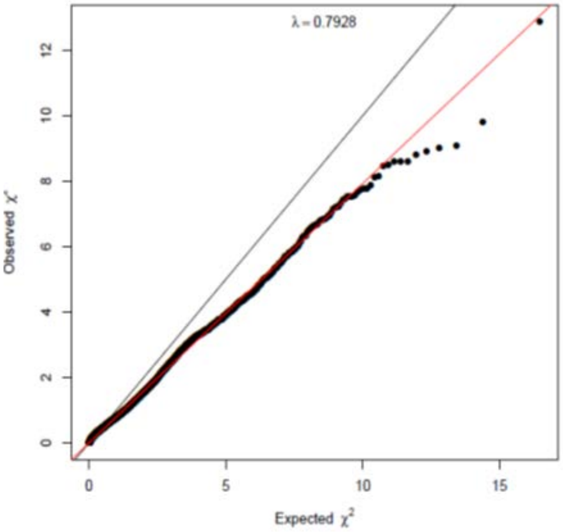

***Supplementary Figure 6.*** Colocalization plot between locus 8p21.3 associated with the composite cardiovascular endpoint in the CHARM-Preserved study and with results from the cardiomyopathy GWAS from the FinnGen project

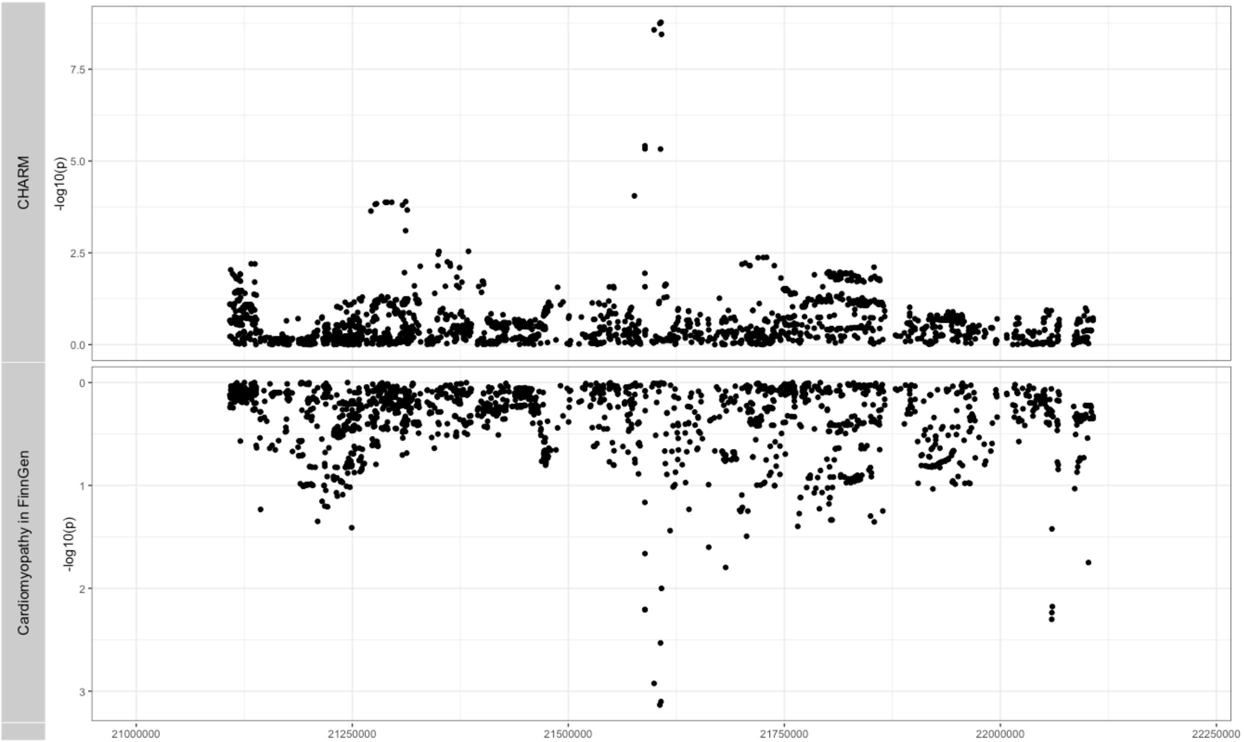

#### References


1. Malik R, Rannikmae K, Traylor M, Georgakis MK, Sargurupremraj M, Markus HS, Hopewell JC, Debette S, Sudlow CLM, Dichgans M, consortium M, the International Stroke Genetics C. **Genome-wide meta-analysis identifies 3 novel loci associated with stroke**. *Ann Neurol. 2018;84(6):934-9.*

2. Nielsen JB, Thorolfsdottir RB, Fritsche LG, Zhou W, Skov MW, Graham SE, Herron TJ, McCarthy S, Schmidt EM, Sveinbjornsson G, Surakka I, Mathis MR, Yamazaki M, Crawford RD, Gabrielsen ME, Skogholt AH, Holmen OL, Lin M, Wolford BN, Dey R, Dalen H, Sulem P, Chung JH, Backman JD, Arnar DO, Thorsteinsdottir U, Baras A, O'Dushlaine C, Holst AG, Wen X, Hornsby W, Dewey FE, Boehnke M, Kheterpal S, Mukherjee B, Lee S, Kang HM, Holm H, Kitzman J, Shavit JA, Jalife J, Brummett CM, Teslovich TM, Carey DJ, Gudbjartsson DF, Stefansson K, Abecasis GR, Hveem K, Willer CJ. **Biobank-driven genomic discovery yields new insight into atrial fibrillation biology**. *Nat Genet. 2018;50(9):1234-9.*

3. Roselli C, Chaffin MD, Weng LC, Aeschbacher S, Ahlberg G, Albert CM, Almgren P, Alonso A, Anderson CD, Aragam KG, Arking DE, Barnard J, Bartz TM, Benjamin EJ, Bihlmeyer NA, Bis JC, Bloom HL, Boerwinkle E, Bottinger EB, Brody JA, Calkins H, Campbell A, Cappola TP, Carlquist J, Chasman DI, Chen LY, Chen YI, Choi EK, Choi SH, Christophersen IE, Chung MK, Cole JW, Conen D, Cook J, Crijns HJ, Cutler MJ, Damrauer SM, Daniels BR, Darbar D, Delgado G, Denny JC, Dichgans M, Dorr M, Dudink EA, Dudley SC, Esa N, Esko T, Eskola M, Fatkin D, Felix SB, Ford I, Franco OH, Geelhoed B, Grewal RP, Gudnason V, Guo X, Gupta N, Gustafsson S, Gutmann R, Hamsten A, Harris TB, Hayward C, Heckbert SR, Hernesniemi J, Hocking LJ, Hofman A, Horimoto A, Huang J, Huang PL, Huffman J, Ingelsson E, Ipek EG, Ito K, Jimenez-Conde J, Johnson R, Jukema JW, Kaab S, Kahonen M, Kamatani Y, Kane JP, Kastrati A, Kathiresan S, Katschnig-Winter P, Kavousi M, Kessler T, Kietselaer BL, Kirchhof P, Kleber ME, Knight S, Krieger JE, Kubo M, Launer LJ, Laurikka J, Lehtimaki T, Leineweber K, Lemaitre RN, Li M, Lim HE, Lin HJ, Lin H, Lind L, Lindgren CM, Lokki ML, London B, Loos RJF, Low SK, Lu Y, Lyytikainen LP, Macfarlane PW, Magnusson PK, Mahajan A, Malik R, Mansur AJ, Marcus GM, Margolin L, Margulies KB, Marz W, McManus DD, Melander O, Mohanty S, Montgomery JA, Morley MP, Morris AP, Muller-Nurasyid M, Natale A, Nazarian S, Neumann B, Newton-Cheh C, Niemeijer MN, Nikus K, Nilsson P, Noordam R, Oellers H, Olesen MS, Orho-Melander M, Padmanabhan S, Pak HN, Pare G, Pedersen NL, Pera J, Pereira A, Porteous D, Psaty BM, Pulit SL, Pullinger CR, Rader DJ, Refsgaard L, Ribases M, Ridker PM, Rienstra M, Risch L, Roden DM, Rosand J, Rosenberg MA, Rost N, Rotter JI, Saba S, Sandhu RK, Schnabel RB, Schramm K, Schunkert H, Schurman C, Scott SA, Seppala I, Shaffer C, Shah S, Shalaby AA, Shim J, Shoemaker MB, Siland JE, Sinisalo J, Sinner MF, Slowik A, Smith AV, Smith BH, Smith JG, Smith JD, Smith NL, Soliman EZ, Sotoodehnia N, Stricker BH, Sun A, Sun H, Svendsen JH, Tanaka T, Tanriverdi K, Taylor KD, Teder-Laving M, Teumer A, Theriault S, Trompet S, Tucker NR, Tveit A, Uitterlinden AG, Van Der Harst P, Van Gelder IC, Van Wagoner DR, Verweij N, Vlachopoulou E, Volker U, Wang B, Weeke PE, Weijs B, Weiss R, Weiss S, Wells QS, Wiggins KL, Wong JA, Woo D, Worrall BB, Yang PS, Yao J, Yoneda ZT, Zeller T, Zeng L, Lubitz SA, Lunetta

KL, Ellinor PT. **Multi-ethnic genome-wide association study for atrial fibrillation**. *Nat Genet. 2018;50(9):1225-33.*

4. Zhou W, Nielsen JB, Fritsche LG, Dey R, Gabrielsen ME, Wolford BN, LeFaive J, VandeHaar P, Gagliano SA, Gifford A, Bastarache LA, Wei WQ, Denny JC, Lin M, Hveem K, Kang HM, Abecasis GR, Willer CJ, Lee S. **Efficiently controlling for case-control imbalance and sample relatedness in large-scale genetic association studies**. *Nat Genet. 2018;50(9):1335-41.*
5. Consortium GT. **The Genotype-Tissue Expression (GTEx) project**. *Nat Genet. 2013;45(6):580-5.*
6. FinnGen. **Documentation of R4 release** *2020. Available from: https://finngen.gitbook.io/documentation/.*
7. Neale_Lab. **UK Biobank GWAS March 2018 resease** *2021. Available from: http://www.nealelab.is/uk-biobank/.*
8. Legault M-A, Perreault L-PL, Dubé M-P. **ExPheWas: a browser for gene-based pheWAS associations**. *medRxiv. 2021:2021.03.17.21253824.*
9. Seifuddin F, Singh K, Suresh A, Judy JT, Chen YC, Chaitankar V, Tunc I, Ruan X, Li P, Chen Y, Cao H, Lee RS, Goes FS, Zandi PP, Jafri MS, Pirooznia M. **lncRNAKB, a knowledgebase of tissue-specific functional annotation and trait association of long noncoding RNA**. *Sci Data. 2020;7(1):326.*
